# Novel brain-penetrant inhibitor of G9a methylase blocks Alzheimer’s disease proteopathology for precision medication

**DOI:** 10.1101/2023.10.25.23297491

**Authors:** Ling Xie, Ryan N. Sheehy, Yan Xiong, Adil Muneer, John A. Wrobel, Kwang-Su Park, Julia Velez, Jing Liu, Yan-Jia Luo, Ya-Dong Li, Luis Quintanilla, Yongyi Li, Chongchong Xu, Mohanish Deshmukh, Zhexing Wen, Jian Jin, Juan Song, Xian Chen

**Affiliations:** Department of Biochemistry & Biophysics, University of North Carolina at Chapel Hill; Chapel Hill, NC 27599, USA; Lineberger Comprehensive Cancer Center, University of North Carolina at Chapel Hill; Chapel Hill, NC 27599, USA; Department of Pharmacology, University of North Carolina at Chapel Hill; Chapel Hill, NC 27599, USA; Neuroscience Center, University of North Carolina at Chapel Hill; Chapel Hill, NC 27599, USA; Mount Sinai Center for Therapeutics Discovery, Departments of Pharmacological Sciences and Oncological Sciences, Tisch Cancer, Institute, Icahn School of Medicine at Mount Sinai; New York, NY 10029, USA; Departments of Psychiatry and Behavioral Sciences, Cell Biology, and Neurology, Emory University School of Medicine; Atlanta, GA 30322, USA; Department of Cell Biology and Physiology, University of North Carolina at Chapel Hill; Chapel Hill, NC 27599, USA

**Keywords:** AD pathogenesis, AD cognitive and noncognitive symptoms, mechanism-based therapy, blood-brain barrier (BBB), brain-penetrant inhibitor, noncanonical translational mechanism, proteomics, proteopathology, mechanism of drug action, signaling pathways, mouse-human conservation, AD diagnosis biomarkers, companion diagnosis

## Abstract

Current amyloid beta-targeting approaches for Alzheimer’s disease (AD) therapeutics only slow cognitive decline for small numbers of patients. This limited efficacy exists because AD is a multifactorial disease whose pathological mechanism(s) and diagnostic biomarkers are largely unknown. Here we report a new mechanism of AD pathogenesis in which the histone methyltransferase G9a noncanonically regulates translation of a hippocampal proteome that defines the proteopathic nature of AD. Accordingly, we developed a novel brain-penetrant inhibitor of G9a, MS1262, across the blood-brain barrier to block this G9a-regulated, proteopathologic mechanism. Intermittent MS1262 treatment of multiple AD mouse models consistently restored both cognitive and noncognitive functions to healthy levels. Comparison of proteomic/phosphoproteomic analyses of MS1262-treated AD mice with human AD patient data identified multiple pathological brain pathways that elaborate amyloid beta and neurofibrillary tangles as well as blood coagulation, from which biomarkers of early stage of AD including SMOC1 were found to be affected by MS1262 treatment. Notably, these results indicated that MS1262 treatment may reduce or avoid the risk of blood clot burst for brain bleeding or a stroke. This mouse-to-human conservation of G9a-translated AD proteopathology suggests that the global, multifaceted effects of MS1262 in mice could extend to relieve all symptoms of AD patients with minimum side effect. In addition, our mechanistically derived biomarkers can be used for stage-specific AD diagnosis and companion diagnosis of individualized drug effects.

**One-Sentence Summary:** A brain-penetrant inhibitor of G9a methylase blocks G9a translational mechanism to reverse Alzheimer’s disease related proteome for effective therapy.

## INTRODUCTION

Alzheimer’s disease (AD), the most common form of dementia in older adults, is a neurodegenerative disorder characterized by progressive decline in cognition, memory, and emotional states^1, 2^. The heterogeneity and multifaceted nature of AD prevents clear mechanistic understanding of disease pathogenesis, which hinders precision diagnosis of AD at a stage when intervention could be effective. The amyloid hypothesis has guided current approaches to AD therapeutics. That is, investigators have developed monoclonal antibody drugs that target amyloid beta oligomers (AβO), such as aducanumab (Aduhelm; Biogen), lecanemab, and donanemab, to reduce Aβ plaques that were postulated to contribute to neuropathologic processes in patient brains^3^. However, these AβO-targeting drugs showed limited specificity and efficacy toward AD in the clinic because (i) the primary drug effect is slowing cognitive decline at the earliest AD stages, but there are no biomarkers to conclusively diagnose early-stage AD, (ii) drug effects were determined primarily based on dose-dependent reductions in plaques measured by positron emission tomography (PET), yet large amounts of amyloid plaques are apparently present in healthy, non-demented individuals^4^, and (iii) no biomarkers other than Aβ plaque size are available to measure effects of these drugs on AD progression, and drug effects on AD biomarkers downstream of Aβ were mixed. Most importantly, multiple pathological brain alterations other than formation of Aβ plaque and neurofibrillary tangles (NFTs) occur during AD pathogenesis, most of which are not readily measurable due to lack of corresponding biomarkers^5^. Because AD patients have highly complicated cognitive and noncognitive symptoms, new mechanism-based drugs are urgently needed for both precision medication of AD and for derivation of new biomarkers to precisely assess drug effects.

Despite the identification of a few AD genetic risk factors^6^, the exact etiology of AD pathogenesis is obscure. Epigenetic mechanisms have been implicated in causing AD pathogenicity because mutations in major histone-modifying enzymes or regulators were characterized in association with AD-related neurodegenerative processes such as impaired memory formation^7^. The histone methyltransferases G9a (EHMT2) and G9a-like protein GLP (hereafter G9a will represent both proteins in their functional dimerized form)^8, 9^ were among the histone-modifying enzymes associated with behavioral abnormalities^10^. Elevated activity of G9a existed in post-mortem brain tissues from AD patients and the familial AD (5xFAD) mice at the AD stage^8^, which implicated G9a activity in AD pathogenesis. In AD, G9a functioned as an epigenetic (transcriptional) suppressor by catalyzing the dimethylation of lysine 9 of histone 3 at specific genes associated with synaptic transmission such as glutamate receptor genes^8^. Recently, Johnson et al. revealed that AD has a proteopathic nature or is a proteomic disease^11^, i.e., AD pathology and cognitive decline strongly correlated with altered expression of a broad spectrum of proteins. However, the canonical, gene-specific transcriptional silencing function of G9a did not explain how expression or modification of specific proteins is regulated in AD conditions in which G9a was constitutively active. That is, the G9a-regulated mechanism responsible for AD-related proteomic alterations (‘proteopathology’) remained to be established.

To determine the function(s) of G9a directly associated with AD pathogenesis, we used our chromatin activity-based chemoproteomics (ChaC)-mass spectrometry (MS)^12^ approach with UNC0965, a biotinylated version of a G9a inhibitor^13^, to dissect G9a pathways in AD. The affinity-tagged inhibitor, which binds only the constitutively active form of G9a, captured proteins that interacted with G9a in AD, and the identities of these proteins were used to deduce G9a-associated pathways. Unexpectedly, ChaC-MS identified a set of translational or post-translational (proteostasis) regulatory proteins that had enhanced interaction with active G9a in AD-related samples. Importantly, the same set of G9a interactors was identified in all AD-related samples including the hippocampus of 5xFAD mice and cerebral organoids derived from AD patients. Particularly, we detected increased interaction between G9a and several regulators of N6-methyladenosine (m^6^A) modification, including METTL3, an RNA methyltransferase that catalyzes m^6^A modification of RNA to promote oncogene translation ^14^; altered expression of METTL3 and m^6^A mRNA were implicated in AD^15, 16, 17^. Recently, our mechanistic study of inflammation control revealed that constitutively active G9a is the upstream regulator of METTL3/m^6^A-mediated translation in chronically inflamed macrophages that mimic AD-related neuroinflammation ^18^. In agreement with findings that translational regulation was broadly perturbed in various neurodegenerative diseases,^19, 20, 21^ and loss of proteostasis is an AD hallmark,^22, 23^ our *in vivo* ChaC-MS identification of G9a interactions with translation regulators such as METTL3 revealed a new, noncanonical function of G9a in the translational or post-translational regulation of AD pathogenesis. Because translational mechanisms ultimately determine function-related protein abundance or modification states^24^, we developed a new mechanism-based therapeutic for AD, i.e., MS1262, a brain-penetrant inhibitor of G9a, to block G9a-mediated translation of AD-related proteins.

The blood-brain barrier (BBB) prevents many drugs from entering the brain; thus, the BBB presents challenges for effective AD therapeutic development^25^. Our new G9a inhibitor MS1262 freely enters the brain. In addition, MS1262 has greater potency than any existing G9a inhibitors (e.g., BIX-01294, UNC0638, UNC0642). We studied the MS1262 effects on two AD mouse models, the 5XFAD and the APP^NLGF^ knock-in (KI)^26^ mice, which are clinically relevant mouse models of AD that comprehensively recapitulate major pathological features of AD patients. Consistently, we found that intermittent MS1262 treatment of both types of AD mice not only fully restored cognitive functions to healthy levels but also reduced anxiety- and depressive-like behavior, typical noncognitive (affective) symptoms of AD patients.

Meanwhile, to discover biomarkers that can be used to 1) precisely diagnose AD patients at early stage, 2) evaluate individual patient response to MS1262 treatment, and 3) stratify appropriate patients for MS1262 treatment with optimized response, we conducted quantitative proteomic and phosphoproteomic experiments^27^ on the hippocampus from the same 5xFAD and APP^NLGF^ KI mouse cohorts that exhibited behavioral improvement after intermittent MS1262 treatment. These AD-correlated proteomic analyses have unique strengths in systematic elucidation of G9a-mediated mechanisms of AD pathogenesis and MS1262 drug action, including drug-affected pathways from which we can derive new AD-specific biomarkers of drug effects. Technically, these unique strengths arise by the genome-wide linking of AD-disturbed, MS1262-recovered protein and phosphorylation levels to specific biological processes/pathways and annotated functional outcomes. As results, our proteomic data indicated that MS1262 treatment reversed or recovered the G9a-regulated, pathogenic expression or phosphorylation of specific proteins that represented AD-disturbed pathways related to learning and memory, cognition, neuronal functions, and non-cognitive behavior. Importantly, these pathway results systematically revealed that AD-activated G9a broadly defines the proteopathic nature of AD.

Presently, an absence of clinically validated protein makers for AD diagnosis prevents validation of pre-clinical results (e.g., drug effects) from animal studies. To overcome this challenge, we systematically determined the clinicopathologic accuracy of MS1262 therapy of AD mice by comparing the AD mouse proteome with AD patient proteomes of statistical significance (> 1000 biospecimen)^11^ This proteome-wide cross-species validation is obviously superior to immunoblotting one protein at a time. Importantly, numerous AD mouse proteins that exhibited G9a/AD-coregulated, MS1262-reversed expression or phosphorylation were components of protein co-expression modules in AD patients^11^ that strongly correlated with human AD pathology and cognitive decline. Specifically, MS1262 treatment reversed the expression and phosphorylation of SMOC1 in the Aβ-associated matrisome module; SMOC1 was found elevated in AD CSF nearly 30 years before the onset of symptoms^28^. Thus, our AD-correlated proteomics identified new biomarkers of early-stage AD and companion diagnosis of individual patient response to MS1262 treatment. In addition, in the clinic, the MS1262 reversal of this mouse-to-human conserved AD proteopathology suggests a G9a-target therapeutic effect for AD patients. Importantly, at the molecular level these proteomic-identified biomarkers revealed that MS1262 treatment broadly alleviated not only cognitive impairments (cognition decline, memory loss) but also lessened noncognitive symptoms associated with AD such as mood disturbances (depression, anxiety). Meanwhile, MS1262 suppressed AD-characteristic expression/phosphorylation of proteins associated with blood coagulation, which may avoid the side effect of brain bleeding or a stroke that were reported for a few AD patients treated by Aduhelm.

In sum, our mechanistic findings show that G9a regulates translation or post-translational modifications (phosphorylation) of a broad range of proteins that are the primary AD executors. Thus, in contrast to existing AD therapies that only slow one symptom (e.g., cognitive decline) targeting AD-activated G9a by the brain-penetrant drug MS1262 showed multifaceted effects that relieve all AD symptoms with minimum side effect. In addition, our AD pathology-correlated proteomic data mechanistically derived multiple biomarkers of early-stage AD and companion diagnosis for individualized, precision medication of AD; some of these biomarkers exist in body fluids accessible for non-invasive diagnosis, which will greatly reduce the burden of patient screening.

## RESULTS

In our overall design (**Fig. 1A**), *in vivo* ChaC-MS dissection of AD brain tissues revealed a previously unknown G9a-mediated translational mechanism amenable to directly limit or reverse AD progression or pathogenesis. The 5xFAD or APP^NLGF^ KI mice that were treated with the brain-penetrant inhibitor MS1262 were used to correlate behavior/synaptic function with analyses of the hippocampus proteome and phosphoproteome. In addition, we compared the proteomic analyses of MS1262-treated AD mice with human AD patient data of statistical significance to identify an AD-characteristic mouse-to-human conserved proteome or phosphoproteome that showed MS1262-reversed expression or phosphorylation. The identification of this AD-correlated proteome/phosphoproteome enabled us to ascertain both G9a translational regulatory pathways associated with AD pathogenesis and the mechanism of MS1262 action on AD pathogenesis. Consequently, we derived new protein biomarkers to evaluate the drug effects on individual patients for precision medication of AD.

**Fig.1.**
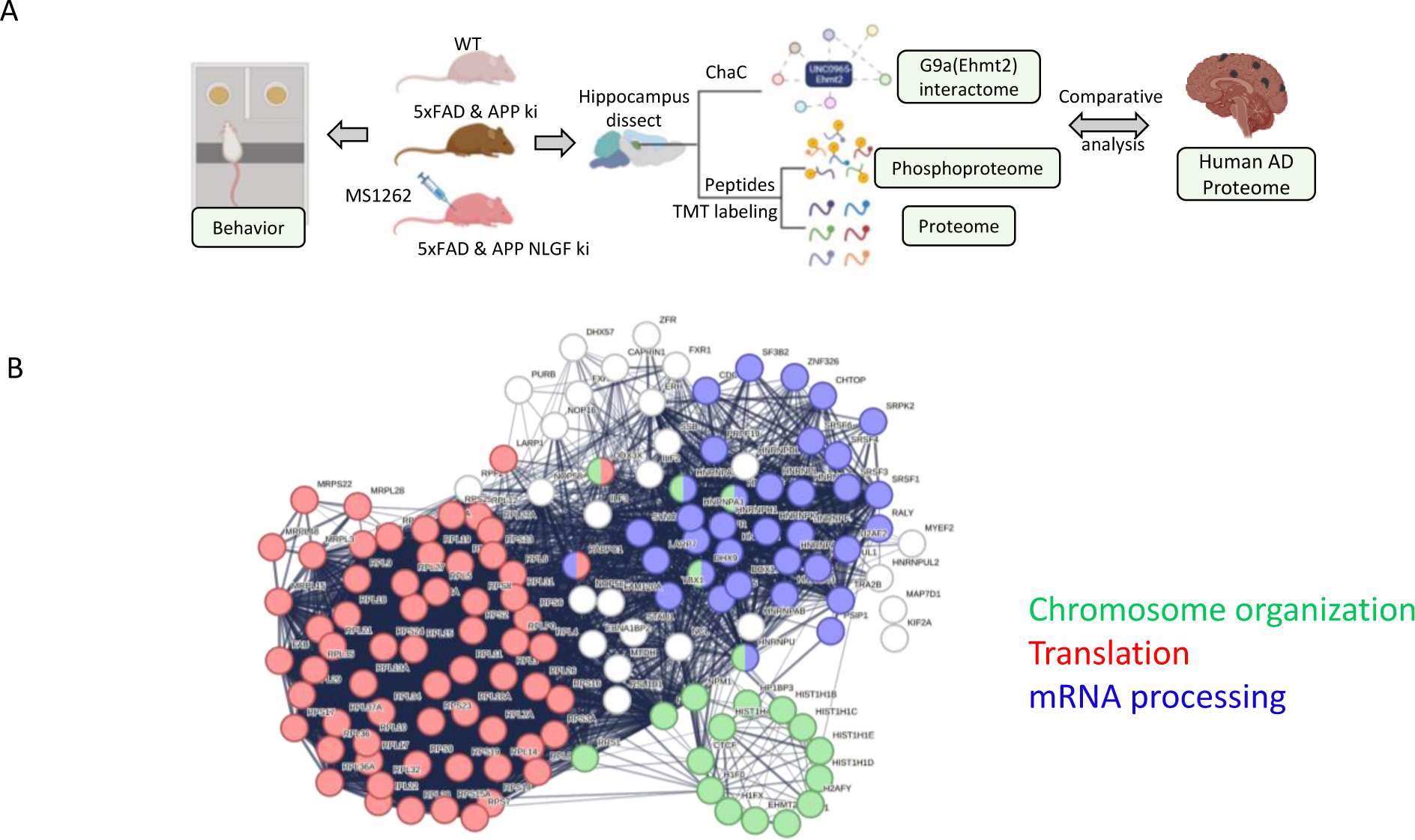
G9a translational mechanism is the target for effective AD therapeutics. (**A**). The overall design for deciphering G9a-mediated AD pathogenesis and the mechanism of G9a-target drug action. (**B**). ChaC-MS revealed a noncanonical function of G9a in the translational regulation of AD pathogenesis. The networks shared by 139 G9a (EHMT2) interactors that were identified in at least 3 of 4 ChaC experiments from 5xFAD mouse hippocampus and AD organoids (human).

### Constitutively active G9a regulates the translational mechanism of AD pathogenesis

We used the biotin-tagged G9a inhibitor UNC0965 with label-free quantitation (LFQ)^27, 29^ ChaC-MS^13^ to identify G9a-interacting proteins in the hippocampus of the 5xFAD mice and cerebral organoids derived from an AD iPSC line F033K with the amyloid precursor protein (APP) V717I mutation^30^. The samples from 5xFAD and normal mice were collected at different ages. The organoid groups at 88 and 102 days represented early AD (MCI) pathology. Based on LFQ ratios (log_2_ > 1, t-test:-log p value > 1.3) that are proportional to the relative binding of individual proteins to G9a, UNC0965 ChaC identified ∼560 proteins that had enhanced interaction with G9a in the hippocampus of AD mice or MCI/AD organoids. Principal component analysis identified a few clusters of G9a interactors, including clusters of AD/MCI-phenotypic G9a interactors, which were well separated from G9a interactors in age-matched, healthy mice or organoids (**fig. S1 and Data S1A & S1B**). These results indicated the high specificity of UNC0965 ChaC-MS to dissect AD heterogeneity with mixed cell types, i.e., UNC0965 captured G9a interactors specifically from AD-related cells that had aberrant G9a activity.

Unexpectedly, the predominant functional networks (mapped by STRING)^31^ overrepresented by the aforementioned AD-phenotypic G9a interactors were primarily associated with major translational or post-translational processes such as alternative splicing, RNA modification and processing, translation initiation and elongation, ribosome biogenesis, and protein degradation (proteostasis^22^) (**Fig. 1B**). Notably, this characteristic profile of G9a-interacting translational regulators was shared between the hippocampus of 5xFAD mice and human AD organoids. Particularly, UNC0965 ChaC-MS identified most known cofactors of METTL3, including HNRNPA2B1, which showed enhanced interaction with G9a in AD-related samples. Jiang et al. reported that progression of tauopathy was mediated by interaction of tau with HNRNPA2B1 and m^6^A RNA ^32^. Similarly, the ChaC-identified interactions of G9a with the METTL3-HNRNPA2B1 translation machinery^14^ implicated a function of G9a in translation associated with tauopathy or AD pathology. More broadly, combined ChaC-MS data predicted that, via AD-phenotypic interactions with key translation regulators such as HNRNPA2B1 and other regulators of ribosomal biogenesis, G9a has a noncanonical (nonepigenetic) function in translational and post-translational regulation of AD pathogenesis.

Also, to determine the clinicopathological relevance of these ChaC findings, we used our multiomics approach (iC-MAP, interaction <u>C</u>orrelated <u>M</u>ulti-omic <u>A</u>berration <u>P</u>atterning)^13^ to retrospectively analyze the NIH Gene Expression Omnibus databases of AD patient blood (Accession: GSE63060)^33^. We identified 26 and 47 G9a interactor mRNAs that had interaction-correlated overexpression patterns^34^ in the blood of 80 MCI and 145 AD patients, respectively, compared with a healthy population (n=104) (**fig. S2**). Interestingly, 20 interactor genes had similarly elevated mRNAs in the peripheral blood cells of both MCI and AD patients^35^ ^36^. This finding revealed a mouse-to-human conservation of AD-related G9a-interacting pathways.

### MS1262 is a highly potent, selective, brain-penetrant inhibitor of G9a and GLP

We previously discovered potent and selective G9a/GLP inhibitors UNC0642 and UNC1479^37^. We and others also reported that the replacement of the quinazoline core with the quinoline core led to higher inhibitory potency toward G9a and GLP, likely due to higher basicity of N-1 of the quinoline core that is crucial for G9a binding^38, 39, 40^. On the basis of these findings, we designed and synthesized MS1262, a novel G9a/GLP inhibitor that contained the quinoline core substituted with a morpholino group at the 2-position (**Fig. 2A**). In biochemical assays, MS1262 (G9a: IC_50_ = 19 ± 14 nM; GLP: IC_50_ = 6 ± 1 nM) showed 23-fold and 37-fold higher potency than UNC0642 (G9a: IC_50_ = 433 ± 6 nM; GLP: IC_50_ = 223 ± 46 nM) for G9a and GLP, respectively (**Fig. 1B**). Using isothermal titration calorimetry, we confirmed that MS1262 had high binding affinities to G9a (*K*_d_ = 74 ± 10 nM) and GLP (*K*_d_ = 19 ± 5 nM) (**Fig. 1C**); these affinities were approximately 3-fold higher than the binding affinities of UNC0642 (G9a: *K*_d_ = 230 ± 17 nM, GLP: *K*_d_ = 62 ± 16 nM)^40^. Moreover, MS1262 displayed excellent selectivity for G9a and GLP, lacking significant inhibition (<20% at 1 µM) for any of the 21 other methyltransferases tested (**Fig. 1D**). Similarly, MS1262 effectively reduced histone H3 lysine 9 dimethylation in cells in a concentration-dependent manner, indicating the strong intracellular inhibitory activity of MS1262 **(Fig. 1E)**. Importantly, MS1262 showed good brain penetration in an in vivo mouse pharmacokinetic study. Following a single 5 mg/kg intraperitoneal (i.p.) injection of MS1262 in C57BL/6 mice, MS1262 displayed brain/plasma ratios of 0.64 and 0.73 at 2 and 4 hours postinjection (**Fig. 1F**). Taken together, these results indicate that MS1262 is a highly potent, selective, brain penetrant G9a/GLP inhibitor suitable for in vivo efficacy studies in AD mouse models.

**Fig. 2.**
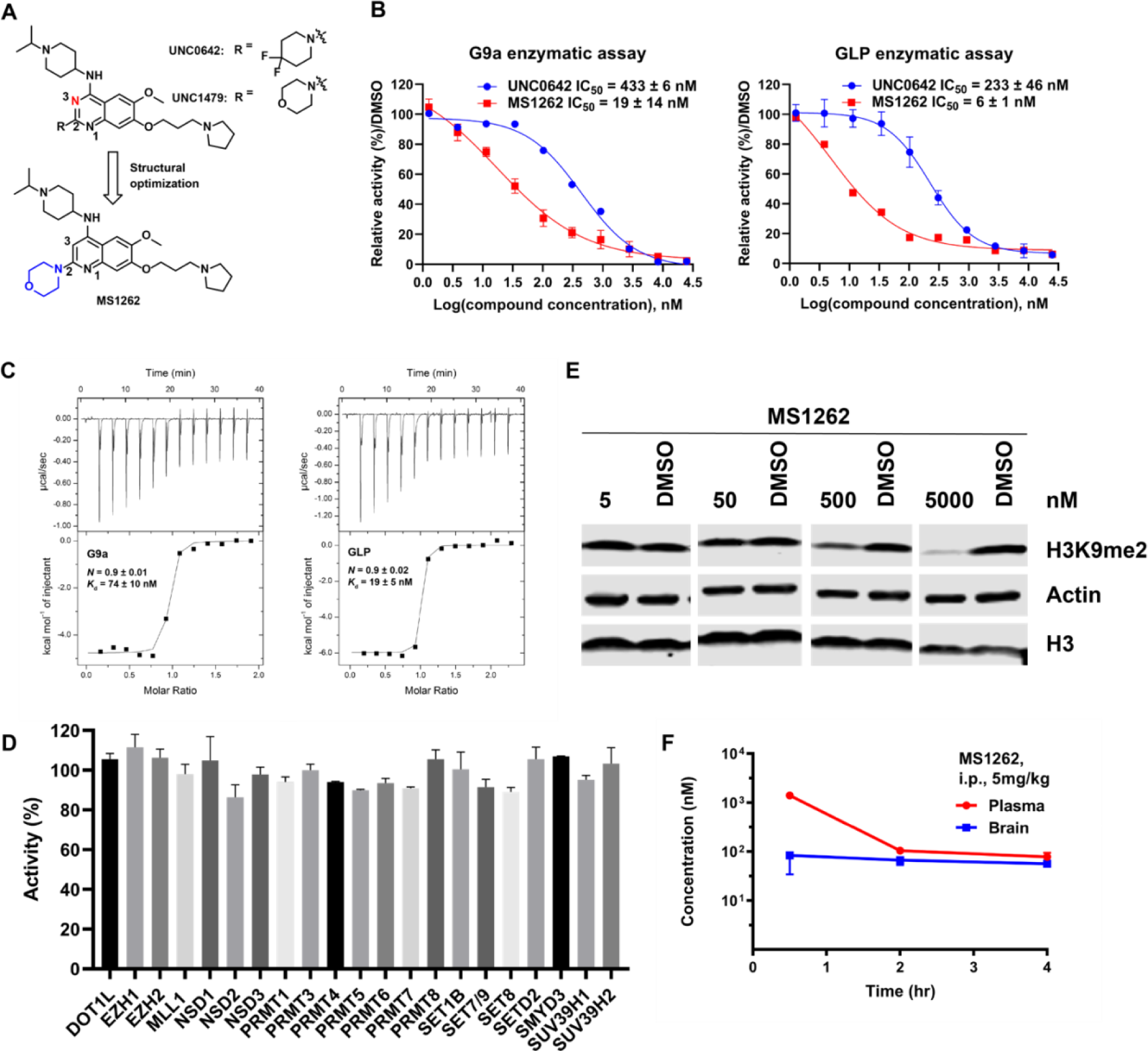
MS1262 is a highly potent, selective, and brain-penetrant inhibitor of G9a and GLP. (**A**). Discovery of MS1262. (**B**). Concentration-dependent inhibition of G9a (top) and GLP (bottom) by MS1262 in G9a and GLP enzymatic assays. UNC0642 was used as a control. Data shown are the mean ± SD from four independent experiments. (**C**). ITC titrations of MS1262 into G9a (top) and GLP (bottom). The calculated values represent the means ± SD from two independent experiments. (**D**). Activity of MS1262 against 21 other methyltransferases at 1 µM. Data are the means ± SD from two duplicate experiments. (**E**). Concentration-dependent reduction of the H3K9me2 level by MS1262 in K562 cells. K562 cells were treated with MS1262 at the indicated concentrations for 48 h. Western blot results are representative from at least two independent experiments. (**F**). Plasma and brain concentrations of MS1262 over 4 h following a single 5 mg/kg intraperitoneal injection of MS1262 in mice. Data shown are the mean ± SD from three tested mice per time point.

### MS1262 inhibition of G9a activity thoroughly rescues AD-related deficits in behavior and synaptic function

The 5xFAD and APP^NLGF^ mice experience cognitive and affective behavioral deficits that progressively worsen with age^41, 42, 43, 44^. Previously, we showed that 5xFAD mice exhibited a series of hippocampus-dependent cognitive and affective deficits, including impaired spatial memory in the novel place recognition (NPR) test, elevated innate anxiety in the open field and zero maze tests, and increased depression-like behavior in the forced swim test^45^. Therefore, we assessed these behaviors in response to long-term, intermittent G9a inhibition by MS1262 (**Fig. 2, A and B**). Locomotion in an open field was unaltered between wild-type (WT) controls and either 5xFAD or APP^NLGF^ mice in response to MS1262 or vehicle treatment (**Fig. 2, C and H**), indicating that G9a inhibition did not affect locomotion. For the NPR test to assess spatial memory, animals treated with MS1262 showed a significant preference for the object in the novel location measured by the discrimination ratio (5xFAD: p<0.0001, APP^NLGF^: p<0.001), indicative of improved spatial memory (**Fig. 2, D and I**). Notably, the discrimination ratio of MS1262-treated mice was rescued to the level of age-matched WT animals for both AD mouse models (**Fig. 2, D and I**). For open field and zero maze tests to assess anxiety-like behavior, MS1262 treatment showed no effects on the amount of time 5xFAD mice spent in the center of an open field (**Fig. 2E**) but MS1262 treatment increased the time APP^NLGF^ mice spent in the center of the open field as compared with both wildtype and vehicle-treated controls (**Fig. 2J**). Furthermore, the 5xFAD and APP^NLGF^ mice spent significantly less time in open arms of the zero maze compared with wild-type control animals, which was significantly increased by MS1262 treatment (**Fig. 2, F and K**). These results suggest the anxiolytic effects by G9a inhibition in AD mice. For the forced swim test to assess depression-like behavior, MS1262-treated animals demonstrated a significantly lower time of immobility compared with vehicle-treated animals, suggestive of less depression-like behavior (**Fig. 2, G and L**). Together, these results suggest that G9a inhibition effectively rescued both cognitive and affective deficits in two complementary AD mouse models, implying broad applicability of effective AD treatment by MS1262.

**Fig. 3.**
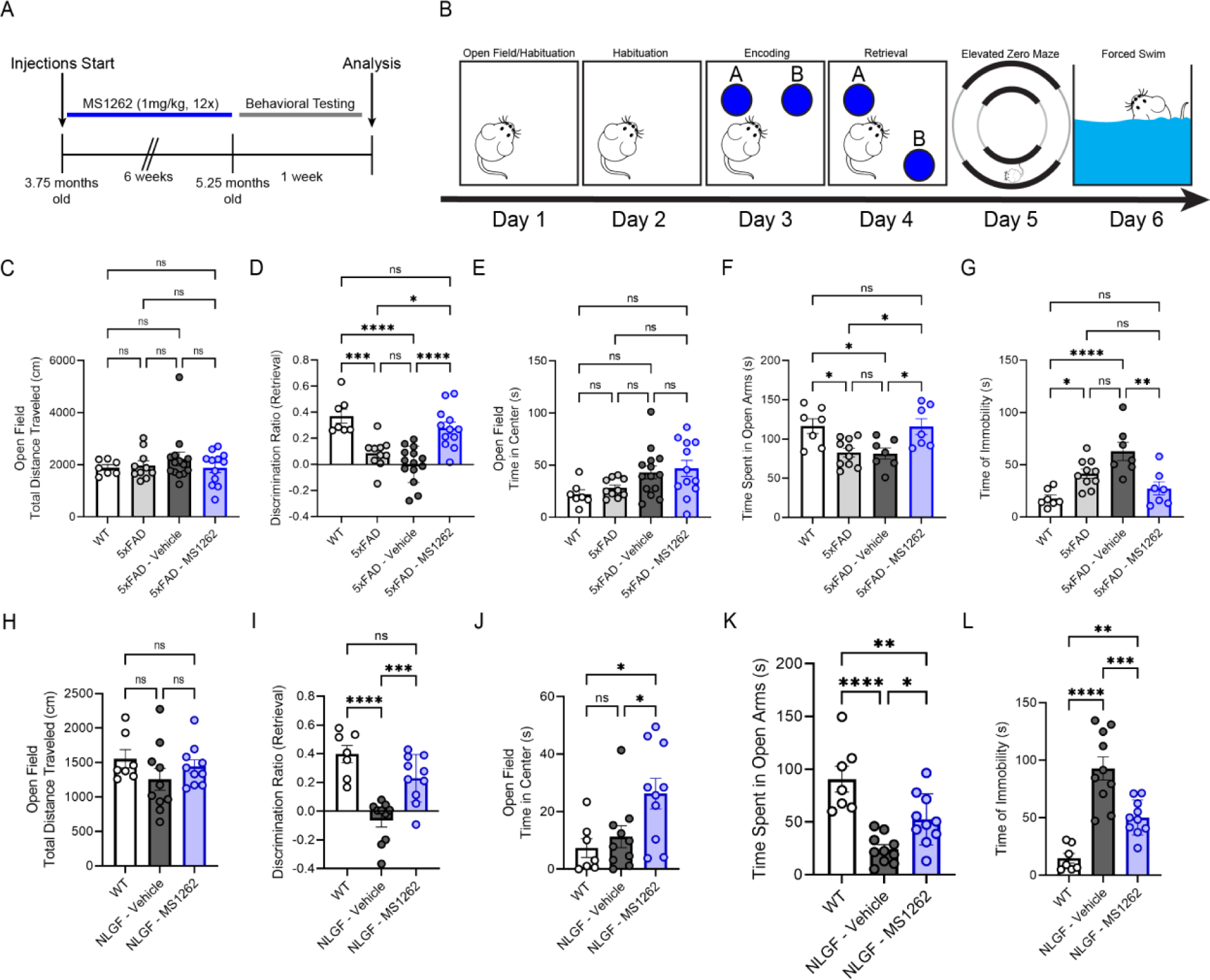
Treatment of 5xFAD mice with MS1262 rescues behavioral deficits. (**A**). Experimental timeline for drug administration and novel place recognition test. (**B**). Depiction of the paradigm used to test memory and affective related behavior. (**C**). Locomotion in an open field was unaffected under wild-type, 5xFAD, vehicle, and chronic MS1262 treatment. (**D**). Preference for the novel-located object during retrieval was significantly reduced in 5xFAD mice compared with wild-type controls and was completely rescued by chronic MS1262 treatment as measured by discrimination ratio (see methods for calculations). (**E**). Time spent in the center of an open field was unaffected under wild-type, 5xFAD, vehicle, and chronic MS1262. (**F**). Chronic MS1262 administration rescued anxiety-like behavior in 5xFAD mice back to wild-type levels demonstrated by increased time spent in the open arms of a zero maze. (**G**). Depressive-like behavioral deficits in 5xFAD mice were rescued to wildtype levels after chronic MS1262 treatment as measured by immobile time during the forced swimming paradigm. (**H**). Locomotion in an open field was unaffected under wildtype, APP^NLGF^, vehicle, and chronic MS1262 treatment. (**I**). Preference for the novel-located object during retrieval was significantly reduced in APP^NLGF^ mice compared to wildtype controls and was rescued by chronic MS1262 treatment as measured by discrimination ratio. (**J**). Time spent in the center of an open field was unaffected under wildtype, APP^NLGF^, while chronic MS1262 treatment led to an increase in time spent in the center. (**K**). Chronic MS1262 administration rescued anxiety-like behavior in APP^NLGF^ mice back to wildtype levels demonstrated by increased time spent in the open arms of a zero maze. (**I**). Depressive-like behavioral deficits in APP^NLGF^ mice were rescued to wildtype levels after chronic MS1262 treatment as measured by immobile time during the forced swimming paradigm. Data are visualized as mean +/-standard error of the mean with each individual displayed as a point. Wildtype (C57BL/6J) (n=7), 5xFAD (n=10), 5xFAD vehicle (n=14), 5xFAD MS1262 (n=12) and Wildtype (C57BL/6J-A^w-J^/J) (n=7), APP^NLGF^ vehicle (n=10), APP^NLGF^ MS1262 (n=10), mice were utilized for behavioral studies. Significance was assessed by ANOVA and Tukey’s posthoc test between each condition. ns = not significant, *p<0.05, **p<0.01, ***p<0.001, ****p<0.0001.

Additionally, to assess the effect of MS1262 treatment on synaptic transmission and intrinsic properties of hippocampal cells, we recorded evoked action potential and pharmacologically isolated spontaneous excitatory postsynaptic currents (sEPSCs) of hippocampal dentate granule cells (GCs) from vehicle- and MS1262-treated 5xFAD mice. The rationale of selecting dentate gyrus GCs for recording was based upon our previous findings showing that the hippocampus-dependent behaviors mentioned above were dentate gyrus dependent^46^. No significant differences were observed between vehicle and MS1262-treated animals in membrane capacity, input resistance, resting membrane potential, and intrinsic excitability (**Fig. 3, A-D**). Interestingly, the frequency of sEPSCs recorded from dentate GCs in MS1262-treated 5xFAD mice was significantly increased without altering the amplitude of sEPSCs (**Fig. 3, F-I**). These results suggested that long-term G9a inhibition by MS1262 in 5xFAD mice increased excitatory glutamatergic synaptic transmission onto the dentate GCs.

**Fig. 4.**
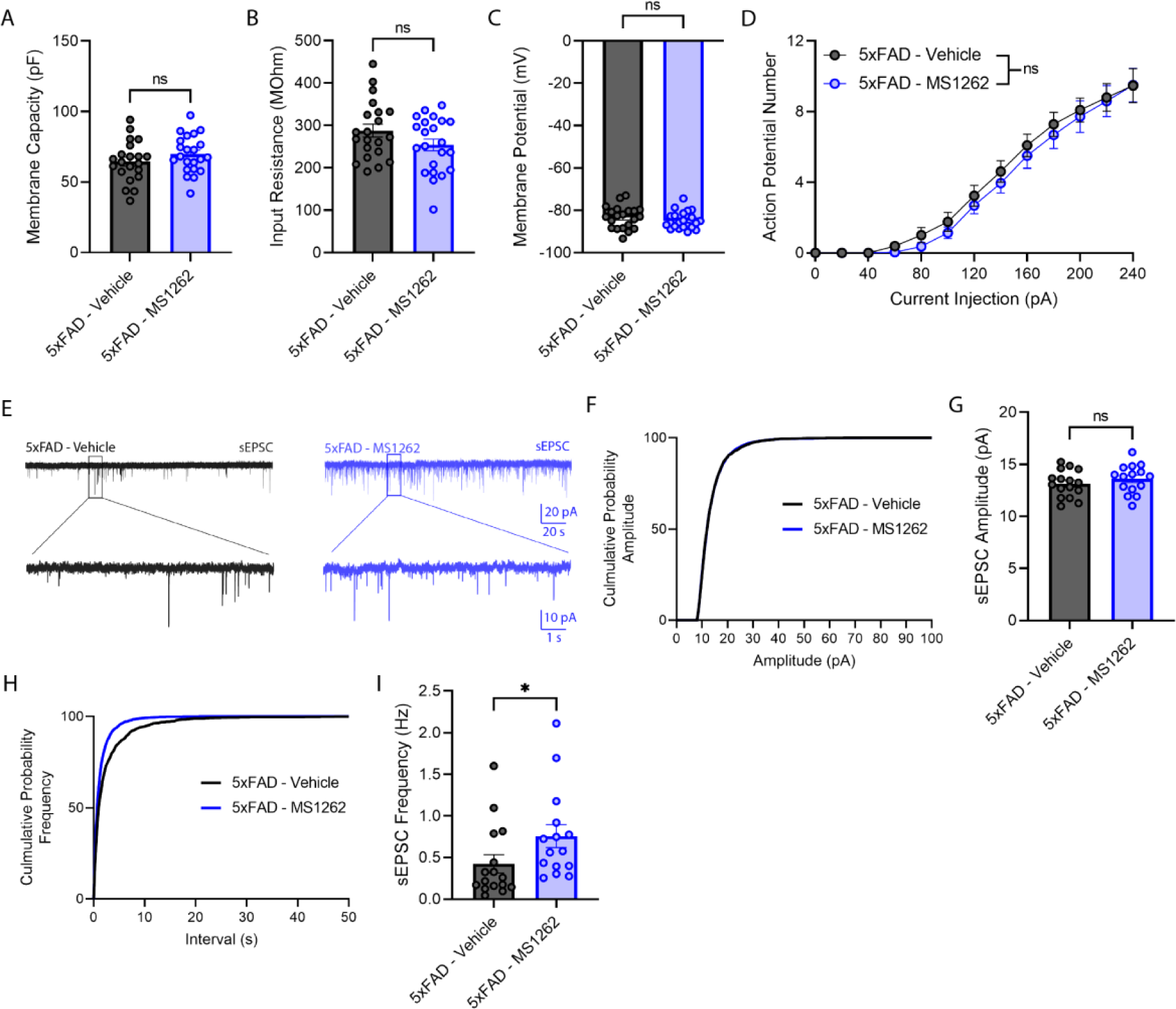
Intermittent MS1262 treatment increases frequency but not amplitude of sEPSCs in DG granule cells of 5xFAD mice without altering intrinsic excitability. (**A**). Quantification of membrane capacity between 5XFAD mice treated with vehicle and MS1262 (n=21/22 cells for vehicle/ MS1262; Two-tailed unpaired Student’s t test, p=0.216). (**B**). Quantification of input resistance between 5XFAD mice treated with vehicle and MS1262 (n=21/22 cells for vehicle/ MS1262; Two-tailed unpaired Student’s t test, P=0.109). (**C**). Quantification of resting membrane potential between 5XFAD mice treated with vehicle and MS1262 (n=21/22 cells for vehicle/ MS1262; Two-tailed unpaired Student’s t test, P=0.311). (**D**). Mean (±SEM) number of action potential elicited in response to increasing step current from dentate granule cells of 5XFAD mice injected with vehicle (black) or MS1262 (blue). (n=21/22 cells for vehicle/ MS1262. Two-way ANOVA: main effect of Interaction, F12,533=0.123, P=0.999. Main effect of Group, F1,533=2.051, P=0.114). (**E**). Representative traces of sEPSCs recorded from dentate granule cells derived from 5XFAD mice injected chronically with vehicle (left) or MS1262 (right). (**F-G**). Cumulative probability (f) and average amplitude distribution (g) of sEPSCs (5xFAD-vehicle, *n* = 16 cells from 3 mice; 5xFAD-MS1262, n = 15 cells from 3 mice. *P* = 0.2995, Mann Whitney test, two-sided). (**H-I**). Cumulative probability distribution (h) and average frequency (i) of sEPSCs (5xFAD-vehicle, *n* = 16 cells from 3 mice; 5xFAD-MS1262, n = 15 cells from 3 mice. *P* = 0.0171, Mann Whitney test, two-sided). \**p* < 0.05, Data are visualized as mean ± SEM.

### A G9a translational mechanism defines the proteopathologic nature of AD

To dissect a G9a-mediated mechanism of AD pathogenesis and the corresponding action mechanism of MS1262 reversal of AD symptoms, we performed ChaC-MS and tandem mass tag (TMT)-based quantitative proteomics/phosphoproteomics experiments ^27, 29^ using microdissected hippocampal samples from the same mouse cohorts used for the aforementioned behavioral studies (**Fig. 2 and Fig. 3**). These samples included AD mice (5XFAD or APP^NLGF^ KI) and age-matched wild-type mice with or without MS1262 treatment. Because the hippocampus is the primary brain area affected by AD, the MS1262-induced changes in G9a binding or changes in the hippocampus proteome/phosphoproteome revealed AD-related, G9a-associated pathways and reflected the inhibitor effects on AD pathogenesis at the molecular level. First, ChaC-MS analysis revealed that MS1262 treatment reversed G9a binding to most regulators of translation, RNA processing, and ribosome biogenesis (correlation coefficient = −0.483, **fig. S3 and Data S1B**). Specifically, MS1262 reversed AD-related binding of G9a to major translation regulators, which confirmed the dependence of G9a translational function on its activity in AD. In addition, in the same G9a-interacting complex in AD, MS1262 inhibition also reversed AD-characteristic G9a interactions with proteins involved in postsynaptic neurotransmitter receptor internalization such as Rabphilin-3A (*Rph3A*), a synaptic vesicle *protein*^47^, methyl CpG binding protein 2 (Mecp2) ^48, 49^, and clathrin-dependent endocytosis such as the hetero-tetrameric assembly protein complex 2 alpha adaptins (AP2A1 and AP2A2)^50^. Thus, combined ChaC results extended G9a function to global translational regulation of AD-related neuropathologic processes.

In parallel, we performed AD-correlated proteomic/phosphoproteomic experiments using micro-dissected hippocampus tissue from a large cohort of 5xFAD mice exhibiting mid/late-stage AD (C57BL/6J; n = 4) with or without MS1262 treatment, along with age matched wild type controls (C57BL/6J; n = 3), resulting in identification of 7,899 proteins and 14,788 phosphorylated sites (in 4,262 protein groups) in total. Similar experiments in a second mouse model of AD, i.e., wild type (C57BL/6J-A^w-J^/J; n = 3) mice versus APP^NLGF^ KI (C57BL/6J-A^w-J^/J; n = 4) mice with or without MS1262 treatment identified 6,576 proteins and 13,6687 phosphorylated sites (in 3,854 protein groups) across all hippocampal samples (**Fig. 4A** and **Data S1C**). Principal component analysis showed not only good separation of wild-type, AD, and MS1262-treated AD 5xFAD/APP-NLFG samples but also showed high quantitative reproducibility between biological replicates (**fig. S4A**). Interestingly, increased m^6^A writer (METTL3, RBM15) and decreased m^6^A eraser (FTO, ALKBH5) expression along with an overall increase in m^6^A modification with age/AD-progression in the hippocampus and cortex of various mouse models (APP/PS1, APPNL-G-F/MAPTP301S) and AD patients has been reported previously ^51, 52, 53, 54^. Similarly, oligomeric tau was shown to complex with m^6^A-modified transcripts through HNRNPA2B1, a m^6^A reader, to regulate translation and promote neurodegeneration ^52^. Interestingly, we recently showed that G9a promotes methyltransferase activity of METTL3 to co-upregulate translation of a subset of m^6^A modified transcripts ^18^, leading us to ask if MS1262 treatment affects m^6^A machinery in AD mouse model. In line with previous reports, we observed increased expression of m^6^A writers (e.g., METTL3, RBM15/15B, WTAP), reduction in m^6^A erasers (e.g., FTO) and dysregulation of several m^6^A readers (e.g., HNRNPA2B1, RBM27, PCIF1, YTHDF2, YTHDC2, HNRNPC/D, etc.) in the hippocampus of 5xFAD and/or APP-NLGF mice, compared to age matched controls; with MS1262 treatment reversing AD-dysregulated expression of several m^6^A regulators (**fig. S4B**). In addition, approximately 47% (5xFAD mice) and 42% (APP-NLGF mice) of proteins that showed AD-characteristic dysregulation that was reversed following MS1262 treatment, classified as ‘AD/G9a-coregulated’ proteins, were translated from m^6^A-tagged RNAs (**fig. S4C**) further confirming the function of enzymatically active G9a in regulating translation of AD-related proteins.

**Fig. 5.**
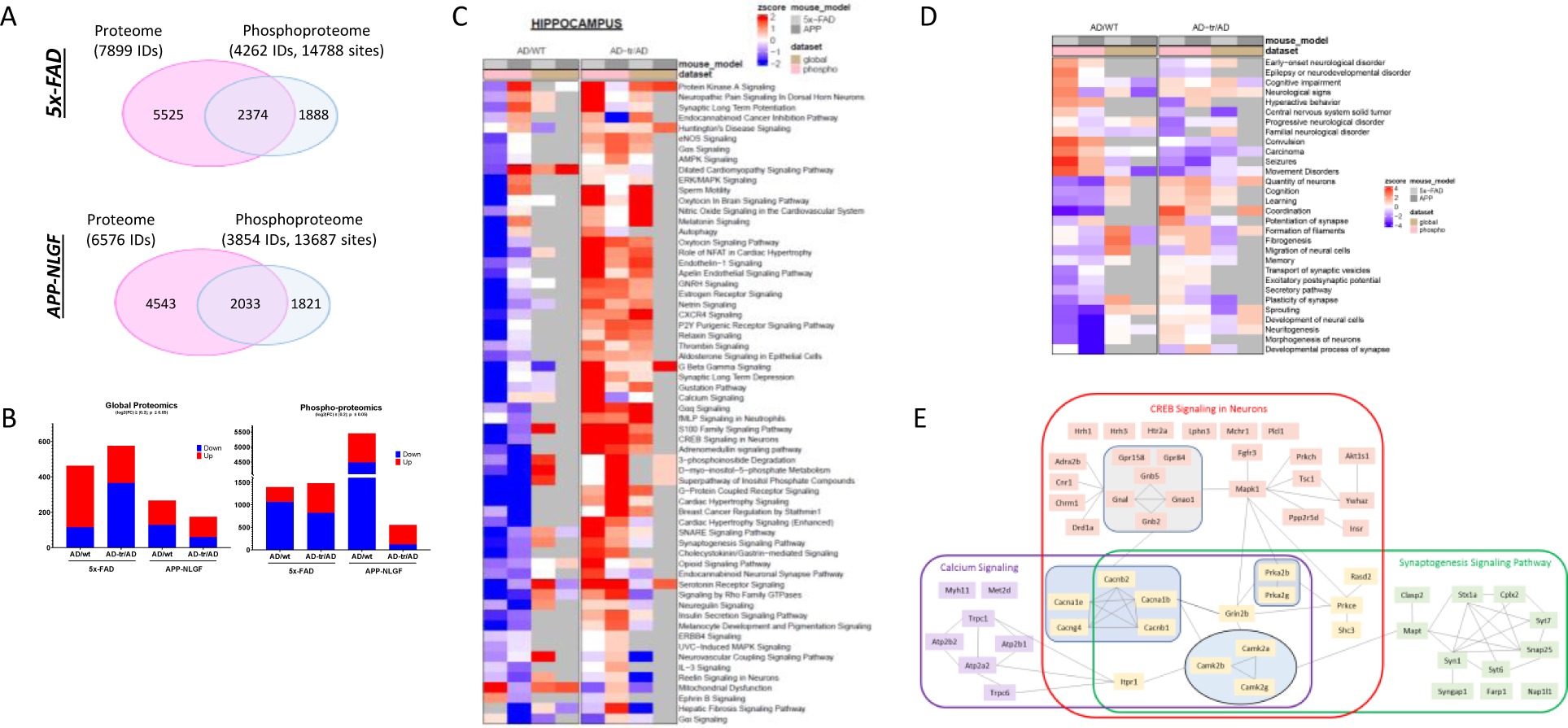
MS1262 inhibition of G9a reversed AD proteopathology (AD-correlated proteome and phosphoproteome). (**A**). Venn diagrams summarizing total number of proteins and phospho-sites identified in 5x-FAD and APP-NLGF mouse models in this study. Briefly, global/phospho-proteomic experiments were performed using proteins extracted from hippocampus of age matched wild-type controls (WT; n = 3), 5x-FAD/APP^NLGF^ mice at mid/late-stage Alzheimer’s (AD; n = 4), and AD mice treated with MS1262 (AD-tr; n =4). Samples were TMT-labeled and run in quadruplicate. (**B**). Bar charts summarizing number of proteins (global proteomics) and phospho-sites (phosphor-proteomics) showing statistically significantly (log2(FC) ± 0.2; p < 0.05; two-tailed unpaired Student’s t test) changes in comparison of AD vs WT and AD-MS1262 treated vs AD mice. **(C).** Heatmap summarizing pathway activity scores, estimated using differentially regulated proteins and phospho-sites shown in (B) for indicated comparisons (AD vs WT & AD-MS1262 treated vs AD) in 5x-FAD and APP-NLFG mice. MS1262 treatment reverses/ameliorates pathway activity changes seen in both mouse models of AD, when compared to age matched wild-type (WT) controls. (**D**). Heatmap summarizing MS1262-affected diseases and functions based on dysregulated proteins/phosphosites shown in (B). (**E**). The network of indicated signaling pathways.

Overall, 9764 entities (global = 1346; phosphosites = 8418; log2FC± 0.2 and P < 0.05) were differentially regulated in our proteomic and phosphoproteomics datasets from 5xFAD and APP-NLGF mice (**Fig. 4B**), with two major clusters of proteins whose expression or phosphorylation levels were either up-or downregulated in 5xFAD and/or APP-NLGF mice compared to age matched controls; a pattern that was reversed following MS1262 treatment (**fig. S4E, and Data S1C**). Because MS1262 specifically inhibited AD-activated G9a, proteins with MS1262-reversed expression or phosphorylation were classified as ‘AD/G9a-coregulated’. Notably, these AD/G9a-coregulated proteins showed more pronounced phosphorylation changes than their expression changes, which confirmed that G9a activity broadly regulates AD pathogenesis at the translational or post-translational (i.e., phosphorylation) levels. Further, we estimated pathway activity scores to find AD-dysregulated (i.e., AD versus wild type) and MS1262-reversed (i.e., treated AD versus AD) biological processes/pathways in the hippocampus of AD mice (**Figs. 5C, fig. S5 and Data S1D**). Notably, most AD/G9a coregulated proteins that were conserved in both 5xFAD and APP^NLGF^ KI mice overrepresented the pathways closely associated with AD pathology or dysregulated neurogenesis. Also, these pathways were associated with indicated functions and diseases (**Fig. 4D, fig. S5B & S5E and Data S1D**). These results confirmed that MS1262 is a proper probe for proteomic discovery of G9a activity-regulated pathways.

As examples, in correlation with MS1262 rescued synaptic functions (**Fig. 3**), MS1262 suppressed AD-characteristic expression of proteins associated with synaptic structure and function, such as neuropilin-1 (NRP1),^55, 56^ tenascin-c (TNC),^57^ and SH3 And Multiple Ankyrin Repeat Domains 3 (SHANK3)^58^ (**fig. S4E**). Also, two of these AD/G9a-coregulated proteins were microtubule-associated protein products of *MAPT* genes, namely, peripheral nervous system Tau (PNS-Tau, UniProt P10637) and Tau-A (or B, C, UniProt B1AQW2) (**fig. S4D**).^59, 60^ Alternative splicing of the *MAPT* transcript produces multiple protein isoforms, and the unique Tau peptide was present in the central nervous system isoforms. We observed that MS1262 treatment suppressed Tau expression that is characteristically increased in AD brains. Because *n*eurofibrillary tangles composed of abnormally hyperphosphorylated Tau are a hallmark of AD,^61^ this result demonstrated the specificity of MS1262 in targeting AD tauopathy.

### G9a-associated pathways mechanistically contribute to both cognitive and non-cognitive symptoms of AD

Broadly on the basis of MS1262-induced phosphorylation changes, we observed that MS1262 reactivated the interactive pathways involved in calcium signaling, CREB (cAMP-response element-binding protein) signaling in neurons, and synaptogenesis signaling (**Fig. 4E**), which are suppressed during AD pathogenesis ^62, 63, 64^. Specifically, SHANK3 is a large scaffolding protein in the postsynaptic density; phosphorylated forms of SHANK3 have different synaptic properties^65, 66^. We found that MS1262 reversed CaMKII (Calcium/CaM-dependent kinase)-mediated phosphorylation of SHANK3^66^ and another synaptic protein GluN2A-subunit-containing NMDAR^67^ (**fig. S5C**) in correlation with improved cognition and rescued synaptic function of MS1262-treated AD mice (**Fig. 3 and 4**). Other AD-suppressed signaling pathways (**Fig. 4C**) were reactivated by MS1262 as described below:

The CDK5 pathway has critical functions in neuronal migration and differentiation, synaptic functions, and memory consolidation. Dysregulated CDK5 activity in AD contributes to formation of senile plaques and neurofibrillary tangles, synaptic damage, mitochondrial impairment of cell cycle reactivation, and neuronal cell apoptosis^68^. In agreement with the observation that silencing CDK5 by RNAi reduced neurofibrillary tangles in the hippocampi of triple-transgenic mice (3xTg-AD mice)^69^. MS1262 treatment protected 5xFAD mice from memory decline and loss of neuronal function presumably by reversing the phosphorylation states of CDK5 pathway proteins (**fig. S5D**).

Dopamine- and cAMP-regulated phosphoprotein (DARPP-32) is a regionally distributed neuronal phosphoprotein that robustly integrates dopamine and glutamate signals in the mammalian brain. cAMP-mediated signaling and cAMP-dependent protein kinase A pathways are crucial in synaptic plasticity and long-term memory^70^. In AD brains, DARPP-32 was cleaved at Thr153 by activated calpain to reduce CREB phosphorylation via loss of its inhibitory function on PP1^71^. MS1262 reversed the phosphorylation-dependent disturbances in CREB function that cause memory deficits in AD patients and animal models^72^. (**Figs. 5C & S5D**)

Dysfunction of the opioid system (opioid receptors and opioid peptides) is implicated in the pathogenesis of AD because of the regulatory functions of opioid receptors in Aβ production, hyperphosphorylated tau, and neuroinflammation^73, 74^. In agreement with the observation that activation of opioid receptors limited the production of Aβ^75, 76^, MS1262 treatment activated the AD-suppressed opioid system by altering the phosphorylation or expression of major components of opioid signaling pathways (**fig. S5D**).

Also, MS1262 reprogramed the regulation of actin-based motility that is associated with cytoskeletal abnormalities and synaptic loss^77, 78^. AD suppression of pathways involving the SNARE complex, oxytocin in spinal neurons, and endocannabinoid system contributes to neurodegeneration and associated impairments in learning, memory, and cognition. The SNARE complex is crucial for vesicle fusion or recycling and neurotransmitter release; inhibition of SNARE complex formation led to neurodegeneration^79^. In correlation with the observation that Injection of oxytocin into the hippocampus reversed some of the damage caused by amyloid plaques in the learning and memory center in an AD animal model^80^, MS1262 treatment reactivated oxytocin in spinal neurons or brain signaling pathways by reversing the phosphorylation states of proteins related to synaptic plasticity and memory formation such as CAMKK1and CAMKK2 ^81^. Also, MS1262 treatment upregulated protein components of brain pyrimidine biosynthesis and pyrimidine salvage pathways that are essential for neuronal membrane generation and maintenance and synapses production^82^, and which were downregulated in AD (**Figs. 5C, 5E & S5**).

Other proteins abnormally expressed in AD and restored to normal levels by MS1262 were gonadotropin releasing hormone, a neuropeptide central regulator of neurogenic and neuroprotective functions and an activator of cAMP-mediated signaling^83, 84^, and G-protein coupled receptors involved in numerous key neurotransmitter systems in the brain that were disrupted in AD ^85, 86^ (**Figs. 5C & S5**). These proteomic/phosphoproteomic results identified multiple signaling pathways that synergistically contribute to *synaptic* plasticity or/and *synaptic* transmission and whose deficits in mice were rescued by MS1262.

In addition, phosphoproteomic analysis identified MS1262-reversed activation states of numerous signaling pathways associated with AD (**Figs. 5C, S5A & S5D)**. Representative pathways include (i) HIF1a signaling related to microglia dysfunction in AD,^87^ (ii) Granulocyte-macrophage colony-stimulating factor with well-established link to AD-related neuroinflammation (astrogliosis),^88^ (iii) Triacylglycerol biosynthesis,^89^ (iv) RHOGDI, Rho-guanosine triphosphatases (GTPases) in regulating the actin cytoskeleton and spine dynamics with the strong association between spine abnormalities (synapse loss) and cognition,^90^ (v) Impaired PI3K-AKT-GSK-3beta-mTOR pathway in AD-associated microglia,^91^ and (vi) Mitochondrial dysfunction^92, 93^. Our phosphoproteomic results indicated that MS1262 treatment simultaneously suppressed these signaling pathways for improved cognition and behavior of AD mice (**fig. S5, D-E**).

In parallel with MS1262-reduced noncognitive AD-like neuropsychiatric behaviors such as depression and anxiety (**Fig. 2**), we identified numerous phosphoproteins with MS1262-reversed phosphorylation involved in Relaxin signaling that contains markers of depression in AD ^94^ (**Fig. 4C & S5D**). Moreover, MS1262 reinstated gustation pathways whose dysregulation was indicative of impaired sensory systems (e.g., olfactory, visual, auditory, somatosensory, gustatory) and poorer memory in AD^95, 96^ because auditory and visual measures were used to detect prodromal AD ^97^ (**fig. S5D**). In sum, the proteins that showed AD-regulated, MS1262-reversed expression or phosphorylation were identified in AD-related, interconnected pathways. These findings demonstrated that G9a is an upstream translation regulator of proteins that define AD proteopathology and associated AD symptoms.

### MS1262 reversed protein expressions or phosphorylation that mark early-stage of AD

To determine the clinical relevance of G9a-translational mechanism that regulates AD proteopathology, and to predict MS1262 efficacy for treating AD patients, we compared our proteomic and phosphoproteomic data from non-treated *versus* MS1262-treated AD mice with the proteomic data of different cohorts of AD patients. We first examined the proteomic data of brain autopsy samples^98^ from (i) control individuals with low pathology of plaques and tangles, (ii) controls with high Aβ pathology but no detectable cognitive defects, (iii) MCI persons with Aβ pathology and slight but measurable defect in cognition, and (iv) AD patients. As shown in **Fig. 5A**, the expression of eighteen proteins including MAPT that mark AD patients was reversed in MS1262-treated AD mice. For example, MS1262 treatment reversed expression or phosphorylation of proteins related to neuroinflammation and AD immunity, including interleukin 33 (IL-33), complement C3, and CD109. Reduced IL-33 expression was observed in the brains of AD mice and AD patients. Similar to the effect of IL-33 injection that reversed cognitive deficits in APP/PS1 mice,^99^ MS1262 inhibition of G9a upregulated IL-33 expression. In correlation with tauopathy, elevated levels of complement C3 protein were detected in brains and cerebrospinal fluid from AD patients.^100^ CD109 is a negative regulator of transforming growth factor β receptor that showed reduced functionality in AD ^101^. These results indicated that the broad function of G9a in regulating AD-related neuroinflammation was alleviated by MS1262 inhibition of G9a. Moreover, SQSTM1 is a multifunctional scaffolding protein that has a major function in autophagy and whose upregulation has been found in several neurodegenerative disorders including AD ^102^. PACAP is a protein encoded by the *ADCYAP1* gene. The close relationship between PACAP reduction and the severity of AD pathology suggests that downregulation of PACAP may contribute to AD pathogenesis ^103^. In addition, ADCYAP1 was identified as a diagnostic biomarker of AD with high discriminatory ability (AUC = 0.850) and validated in AD brains (AUC = 0.935). PBXIP1 is a cerebrospinal fluid (CSF) marker of AD. Comparison of cortex and serum led to an AD-correlated protein panel of CTHRC1, GFAP and OLFM3.^104^ The CAMKK2-AMPK kinase pathway mediates the early synaptic toxic effects of Aβ42 oligomers and is a target for AD therapeutics ^105^. In addition, we found that MS1262 reversed phosphorylation of the protein products of 33 AD-risk genes^106^ (**fig. S5C**).

**Fig. 6.**
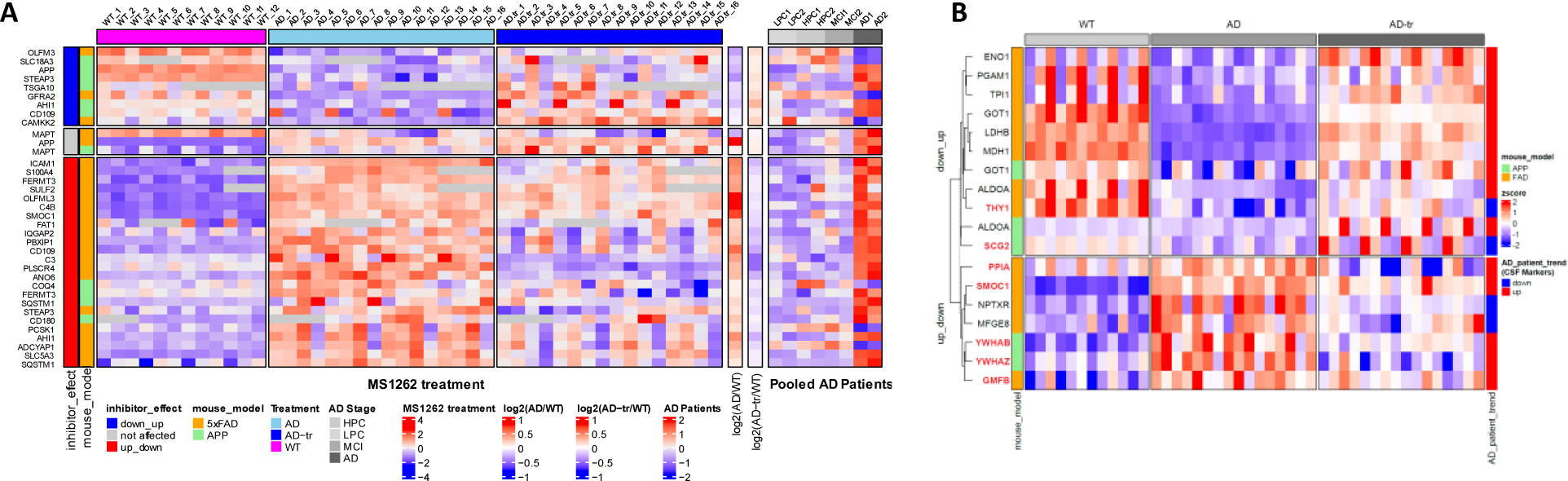
MS1262 treatment reversed expression or phosphorylation of protein markers of AD risk or early AD stage. (**A**) Heatmap of select AD-risk markers showing patient-correlated protein expression in 5x-FAD & APP-NLGF mouse models whose expression was reversed by MS1262 treatment. Global proteomics data from three age matched wild-type controls (WT), four 5x-FAD/APP^NLGF^ mice at mid-/late-stage Alzheimer’s (AD), and four AD mice treated with MS1262 (AD-tr), with each sample run in quadruplicate. Rows are clustered into three groups based on pattern of inhibitor effect: (1) down_up = markers down in AD mice, compared to controls, whose expression is back up following MS1262 treatment, (2) up_down = markers whose expression increases in AD mice, compared to controls, but is back down upon treatment, and (3) not_affected = inhibitor did not affect expression of these markers in AD mice, compared to controls. The left annotation column (mouse_model) shows the proteomics dataset (orange = 5x-FAD, green = APP-NLGF) a marker belongs to. Right annotation columns [log2(AD/WT) & log2(AD-tr/AD)] show fold change for indicated comparisons. Last heatmap on the right shows AD marker expression in the Banner Sun individual cohort for (1) controls with a low pathology of plaques and tangles (LPCs), (2) controls with high Aβ pathology but no detectable cognitive defects (HPCs), (3) mild cognitive impairment (MCI) with Aβ pathology and a slight but measurable defect in cognition and (4) late-stage AD with high pathology scores of plaques and tangles. Pooled samples from a total of 39 Banner Sun individual cases were measured with TMT-LC/LC-MS/MS in the four disease groups (LPC = 12, HPC = 6, MCI = 6, and AD = 15) and are represented by boxes filled with red gradients. FC: fold change. (**B**) MS1262 reversed cerebrospinal fluid (CSF) proteome of early-stage AD. Heatmap showing AD markers (identified from CSF of symptomatic/non-symptomatic AD patients) dysregulated in 5xFAD and APP mice, compared to DMSO treated wild-type controls, whose expression is reversed following MS1262 treatment. The annotation column on right indicates up (red) or down (blue) regulation of said marker in symptomatic AD patients compared to age matched non-symptomatic patients. Proteins names highlighted in red have AD-dysregulated and G9a reversed expression pattern that shows mouse-to-human conservation.

Further, to identify mouse-to-patient conserved biomarkers for precision diagnosis at a stage when MS1262 treatment could be effective, we compared our mouse profiles of G9a/AD-coregulated, MS1262-reversed proteins with CSF protein biomarkers of AD progression ^28^. Notably, MS1262 treatment reversed AD-characteristic expression of multiple CSF biomarkers of early-stage AD (**Fig. 5B**). For example, the level of SPARC-related modular calcium-binding protein 1 (SMOC1), an Aβ plaque-associated synaptic protein^23, 107^ was found elevated in AD CSF nearly 30 years before the onset of symptoms ^28^. Other CSF markers ^28^ showing MS1262 affected/reversed expression included the following (**Fig. 5B**): Glia maturation factor β (GMFB), a newly characterized G9a-regulated factor in the regulation of neuronal and glial growth and differentiation^108^; Thy-1 glycoprotein, which was implicated in extensive growth of abnormal neurites in AD ^109^; proton pump inhibitor A (PPIA), which was related to cognitive decline ^110^; and SCGC, which is typically transported in synaptic vesicles as a marker of synaptic loss and neuronal injury/degeneration for prognosis in prodromal AD ^111^. We also noticed that two CSF markers showed AD-upregulated, MS1262-suppressed expression in the hippocampus of AD mice but decreased abundance in patient CSF, namely, MFGE8, which was implicated in Aβ-induced phagoptosis and a potential therapeutic target to prevent neuronal loss in AD ^112^ and neuronal pentraxin receptor (NPTXR/NPTX2), which was identified as a biomarker of AD progression for CSF-based liquid biopsy ^111, 113^. These discrepancies of AD-related expression were probably due to different sample origins from the hippocampus or CSF, respectively. Nevertheless, simultaneous identification of these CSF biomarkers of AD as MS1262/G9a-regulated proteins suggested that these biomarkers together can be used to stratify appropriate patients who may have maximum response to MS1262 treatment and to determine individual effects of MS1262 treatment.

### MS1262 reversed multiple AD-specific brain pathological processes that mediate Aβ plaque and NFT pathology

We identified and focused on two major clusters of proteins/phospho-proteins whose expression (n = 96) or phosphorylation (n = 252) levels were either up-or downregulated in 5x-FAD and/or APP-NLGF mice compared to age matched controls; a pattern that was reversed following MS1262 treatment (**Fig. 6A, and Data S1C**). These ‘AD/G9a-coregulated’ entities were primarily involved in synaptic signaling, neuronal transmission and nervous system development related pathways (**Fig. 6B**) with 148 out of 348 members showing Aβ peptide (residues 6-28) correlated expression in the brain of AD patients including CD109, CD44, ANO6, ICAM1 and PLSCR4 (**Fig. 6D, fig. S6A and Data S1E**). Inflammation-related CD44 splice variants are overexpressed in AD hippocampus and may protect neurons from AD damage ^114^. TMEM16F (also known as ANO6) mediated microglia polarization is involved in progression of AD and its inhibition shows neuroprotective effects in AD models ^115^. ICAM-1 improves cognitive behaviors in 5xFAD mice by inhibiting NF-κB signaling ^116^ while PLSCR4 is a transcriptome biomarker hub gene in 5xFAD mice ^117^. Overall, these AD/G9a-corregulated proteins were involved in mental/CNS/neurodevelopmental disorders, memory impairment, mental deterioration, and thrombosis/coagulation, among other diseases (**Figs. 7C & S6B**). These results confirmed that G9a activity is closely associated with AD pathology or dysregulated neurogenesis and MS1262 is a proper probe for proteomic discovery of G9a activity-regulated pathways.

**Fig. 7.**
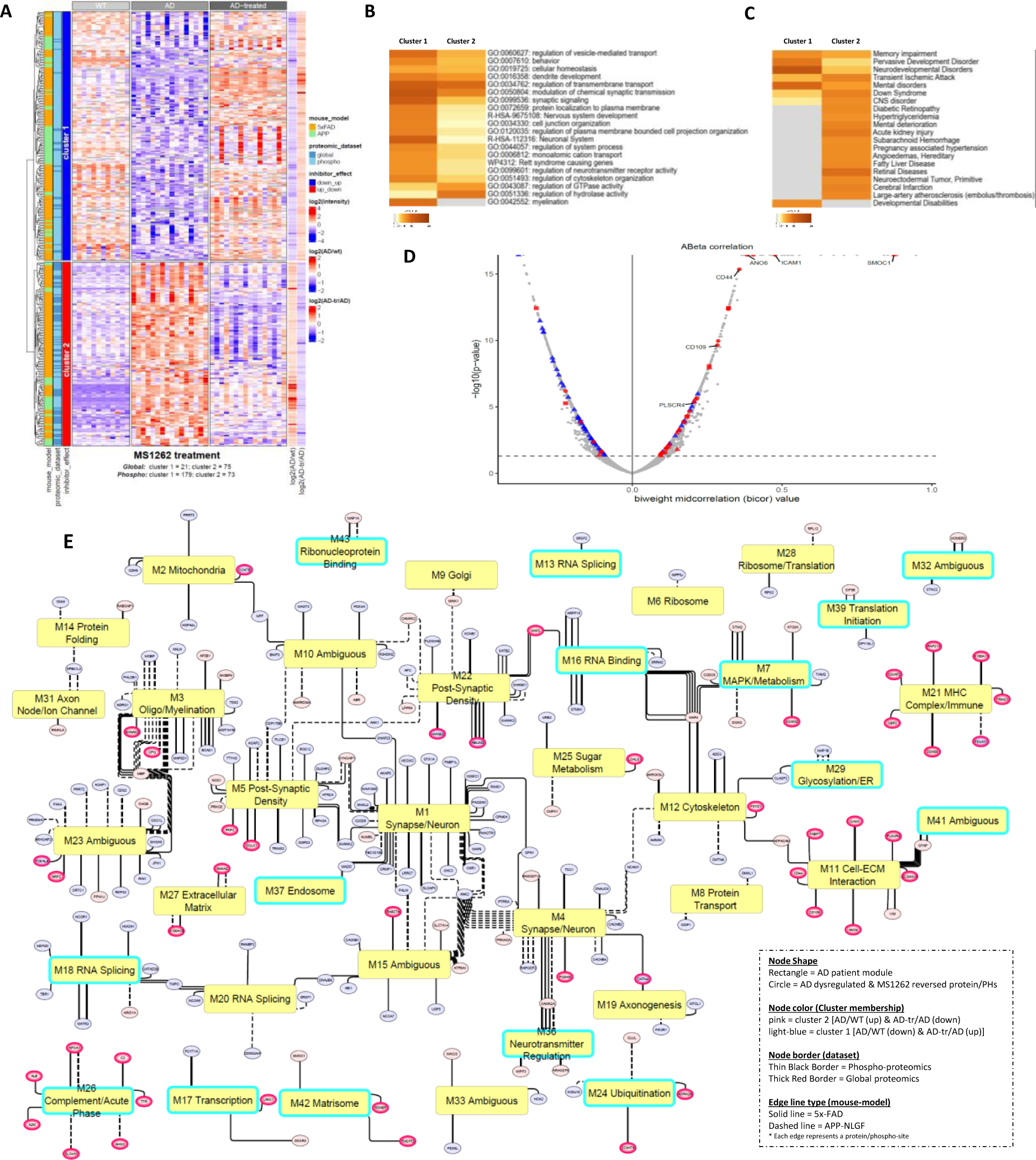
MS1262 treatment of AD mice reversed AD patient proteome or phosphoproteome. **(A)** Heatmap depicting AD/G9a co-regulated proteins/phosphoproteins showing dysregulation in 5x-FAD/APP-NLGF mice, compared to age-matched controls, whose expression/phosphorylation pattern is reversed following MS1262 treatment. Right annotation columns depict log2(fold-change) for indicated comparisons (AD/WT & AD-tr/AD). **(B-C)** Heatmaps summarizing results of GO/pathway enrichment (B) and disease & function (C) analysis for proteins and phospho-proteins belonging to the two clusters identified in (A). **(D)** Volcano plot illustrating the relationship between expression levels of various proteins and the Aβ peptides (residues 6-28) in the brain of AD patients (n = 488)^11^. The plot shows the biweight mid-correlation (bicor) coefficient and the corresponding p-value for each protein. Cluster 1 (blue) and cluster 2 (red) proteins from (A) are highlighted with point shape denoting proteins (circle) and phospho-proteins (triangle). Names of select AD markers are shown. **(E)** Network illustrating the relationship between AD/G9a-corregulated proteins/phosphoproteins identified in (A) and various AD-patient correlated modules (’M’) that are dysregulated in the brains of AD patients ^11^. Modules showing dysregulation at protein level only, without concomitant change at the transcriptional level in AD patients, are highlighted in cyan.

Next, we compared AD/G9a-corregulated proteins identified in 5xFAD/APP-NLGF mice with “modules” (M) of co-expressed proteins showing dysregulation in brains of AD patient ^11^ to identify 204 entities (51 proteins, 204 phosphoproteins) with AD-related, MS1262-reversed expression changes (**Fig. 6E**). Notably, 15 out of 38 of these AD/G9a-corregulated ‘modules’ were only dysregulated at the protein level without concomitant change in transcriptomic networks in the AD patient samples (**Fig. 6E**; highlighted in cyan). This finding, along with the observation that nearly half of the AD/G9a-corregulated proteins are encoded by m^6^A-tagged transcripts (fig. S4C) and the fact that AD/G9a-coregulated proteins exhibited more pronounced phosphorylation changes than expression changes, further confirmed that G9a activity broadly influences AD pathogenesis at the translational or post-translational (i.e., phosphorylation) levels.

In the networks of patient protein modules (**Fig. 6E)**, SMOC1, whose level in CSF or in postmortem brain was associated with AD pathology, was an MS1262-targeted component of the M42 matrisome module strongly correlated with AD neuropathology and cognition. The Tau family microtubule-associated proteins^118^ and STIM2 responsible for neuronal calcium homeostasis impairment^119^ were MS1262-targeted components of module M7 MAPK signaling and metabolism that is highly associated with the rate of cognitive decline. Plasma glial fibrillary acidic protein, an astrocyte reactivity biomarkers for AD^120^ and nitric oxide synthase 1 for synaptic transmission and neuroplasticity^121^ were MS1262-targeted components of modules M5 post-synaptic density or M11 cell-ECM interaction, respectively, again, major modules strongly correlated to AD neuropathology and cognition. In addition, MS1262 reversed the phosphorylation of components of modules M29, glycosylation/ER, and M42, matrisome, that were correlated with AD endophenotypes, and elevated tau microtubule-binding domain peptide levels correlate with the other MS1262-affected components of M42 matrisome and M11 cell-ECM modules. Overall, AD/G9a-corregulated co-expression modules were involved in neuronal signaling (neurotransmitter regulation, axonogenesis, synapse signaling, myelination, postsynaptic density), translation (RNP binding, translation initiation), RNA metabolism (transcription, splicing, binding), protein transport (glycosylation/ER, Golgi, endosome), cellular energetics (sugar metabolism, mitochondria) and immune response (complement/acute phase, MHC complex).

In short, we examined spatial relationships between MS1262-reversed AD patient proteome and Aβ plaques and neurofibrillary tangles (NFTs), the hallmarks of AD neuropathologies. In addition to MS1262 reversal of the phosphorylation of SMOC1 in M42 matrisome, MS1262 reversed the phosphorylation of many module components found in Aβ plaques or/and NFTs, including M1 and M4 components enriched in both Aβ plaques and NFTs, M7 components primarily associated with Aβ plaque, and M13 and M29 components uniquely enriched in NFTs. Notably, the levels of mRNAs encoding these module proteins showed little change in AD patients, although AD-upregulated expression or phosphorylation of these proteins was reversed by MS1262 treatment. This lack of correlation between mRNA and protein expression indicated that constitutively active G9a regulates the translation and post-translational modification (phosphorylation) of AD-related proteins, and these proteins mediate protein co-localization with Aβ plaques and NFTs.

In addition, we found that these G9a-dependent, AD-related protein co-expression and co-phosphorylation modules were preserved across the mouse and human samples as G9a-translated because MS1262-targeted proteomic or phosphoproteomic changes were more closely aligned with AD pathology and AD-like behavior than changes at the mRNA level. In parallel with the observation that MS1262 repaired AD-related behavior, MS1262 treatment reversed the proteopathologic landscape in AD patients, which was evidenced by the greater number of phosphoproteins in the protein co-expression modules highly correlated with broad AD pathology and cognitive deficits. In the clinic, these proteomic identifications of AD mouse-to-patient conservation revealed multiple MS1262-affected biomarkers of AD which can be used to evaluate evidence of downstream disease modification and to assess the relation between biomarker changes and clinical outcomes.

## Discussion

Epigenetic mechanisms involving G9a are thought to regulate AD pathogenesis. However, few pathways strongly correlated with AD pathology were identified by transcriptomic analysis of G9a inhibitor (UNC0642)-treated 5XFAD mice.^8, 122^ Instead, recent multiomics analyses of large cohort samples of AD patients^11^ identified the proteopathologic nature of AD, i.e., proteomic changes that were not observed at the mRNA level strongly correlated with AD pathology and cognitive decline, which suggested that AD pathogenesis is predominately a translation abnormality. Using chemical biology, animal behavioral analysis, and AD-correlated proteomics, we discovered a noncanonical, translation-regulatory function of aberrantly activated G9a that defines the proteopathologic nature of AD. Accordingly, we have developed new mechanism-based, brain-penetrant small molecule therapeutics that inhibit the G9a-mediated translational mechanism in AD.

Present evidence is insufficient to support the hypothesis that abnormal *Aβ deposits* are uniquely causal in AD progression; it is unknown whether a large *Aβ* plaque could be the cause or a result of AD. Also, the Aβ aggregation cascade is composed of a mix of species; thus, any Aβ-targeting monoclonal antibody has limited specificity/efficacy toward AD. In contrast, our AD therapeutics approach is based on a new translation-regulatory mechanism of AD pathogenesis whereby AD-activated G9a translates select mRNAs into proteins that are widespread in a range of pathological brain processes that mediate Aβ plaque and NFT pathology. In correlation with AD behavior reversal, we employed G9a inhibitors (either biotin-tagged or tag-free forms) as the pathway probes for AD-correlated proteomic dissection of G9a-regulated AD pathogenesis and the mechanism of MS1262 drug action. First, ChaC-MS identifed AD-characteristic, MS1262-reversed G9a interactions with proteins that constitute the ‘pathway skeleton’ of the translational control of AD pathogenesis. These ChaC-identified G9a interactors included regulators of either translational processes, such as alternative splicing, ribosome biogenesis, protein synthesis, and proteostasis,^22, 23^ or neuropathogenic processes related to neuronal endocytosis and synaptic functions. These AD-phenotypic G9a-interacting translation regulators include multiple m^6^A RNA regulators such as HNRNPA2B1, YTHDC2, and YTHDF2, all of which were implicated in AD.^32^ Our m^6^A RNA-to-protein correlation analysis identified G9a/AD-coregulated m^6^A mRNAs whose protein translation was reversed by MS1262 treatment of AD mice, validating G9a regulation of m^6^A-mediated translation in AD. In addition to the m^6^A translation regulatory axis, G9a also interacted with other translation regulators. For example, SRRM2 contributes the tauopathies in AD by mislocalizing to cytosolic tau aggregates in brains of individuals with AD.^123^ HNRNPK, a splicing factor, regulates AD-driving proteoforms.^124^ The ribosomal protein interactors RPS14, RPS17, RPS15A, RPL21, RPS8, RPL37A, RPL14, RPS2, RPL26, FAU, RPS13, RPS24 were significantly upregulated in AD patients.^125^ The NFT-associated AP2A1 and AP2A2^50^ were identified as G9a interactors that are the central hub for AD pathogenesis-associated clathrin-dependent endocytosis and postsynaptic neurotransmitter receptor internalization. The synaptic protein *Rph3A* associated with Aβ burden^47^ when Mecp2 was implicated in neuronal endocytosis and synaptic plasticity.^48, 49^ In line with our finding of similar G9a translational function in chronically inflamed macrophages, these results of G9a interactions with translation regulators confirmed that G9a activity is crucial for global translation of AD-causative proteins.

Specifically for AD therapy, our small molecule G9a inhibitor MS1262 showed high brain permeability and stability and much higher potency than existing G9a inhibitors, e.g., > 20-fold more potent than UNC0642 (the older version of G9a inhibitor). As a result, MS1262 inhibition of G9a fully rescued both cognitive and noncognitive behavioral deficits of 5xFAD mice and APP^NLGF^ KI mice (**Fig. 3 and 4**). In a nonbiased manner, our global proteomic profiling revealed that MS1262 systematically reversed the expression or phosphorylation of a range of proteins in all major neurodegenerative or AD-related signaling pathways; little MS1262 effect was observed on expression or phosphorylation of non-AD related proteins in 5xFAD mice. In addition, select mRNAs, termed as ‘poised’ mRNAs, showed little changes but produced specific proteins associated with AD pathogenesis. In patient samples we identified MS1262-regulated, ‘poised’ mRNAs that are translated into AD-driving proteins. These proteomic data demonstrated that MS1262 crossed the BBB directly and specifically targeted G9a-regulated proteomic pathways to reverse AD proteopathy. Notably, not all but most AD-related pathways that are overrepresented by G9a/AD-coregulated proteins were identified in both mouse models. The discrepancy of other pathways is probably due to differences in the background of mice in the wild-type (C57BL/6J) and 5xFAD (C57BL/6J) experiments and the mice of the wild-type (C57BL/6J-A^w-J^/J) and APP^NLGF^ (C57BL/6J-A^w-J^/J). Furthermore, 5.5 months old of 5xFAD mice were considered as late-stage AD^8^, but the same-aged APP^NLGF^ KI mice were at an earlier stage of AD due to the slow progression of AD pathology in the knock-in line^126, 127^.

In addition, in contrast to the fact that MS1262 inhibition of G9a reversed the widespread expression of proteins that are causative in AD pathology, we found that treatment of AD mice by UNC0642 had little effect on the global expression of AD-related proteins in the hippocampus (**data not shown**); this limited effect was probably due to the low brain permeability/stability or relatively low potency of this compound.

Compared with Aβ-targeting monoclonal antibodies that only slow cognitive decline of small numbers (e.g., 27%) of patients at the earliest stage, MS1262 displayed high efficacy and specificity/sensitivity toward AD, i.e., MS1262 showed multifaceted, therapeutic effects on AD including cognition restoration, memory improvement, and attenuation of depression and anxiety. Because AD-activated G9a broadly and simultaneously regulated major signaling pathways that mechanistically contribute to both cognitive and non-cognitive symptoms of AD, MS1262 effects are not ‘one target (G9a) at a time’ but, instead, the drug effect (target) is proteome-wide. As supporting evidence, Pao et al. reported^128^ that targeting CDK5 hyperactivity ameliorated neurodegenerative phenotypes. Accordingly, we identified CDK5 signaling as a one of many G9a/AD-coregulated pathways that were simultaneously reversed by MS1262 treatment. This identification of CDK5 confirmed that G9a is a broad regulator of AD proteopathologic landscape, hence, MS1262 rescued a range of AD-dysregulated pathways that define multiple AD pathological hallmarks such as Aβ plaque and NFT pathology. In addition, AD mice treated with MS1262 for six weeks did not show any signs of toxicity; general appearance, activity, and behavior were all normal or improved. Outward normality was consistent with MS1262 reversal of AD proteome and phosphoproteome of the hippocampus. On a systems evaluation of MS1262 side effects, our AD pathology-correlated proteomic/phosphoproteomic results demonstrated that MS1262 specifically and effectively inhibited G9a that was aberrantly activated in AD-related cells but not non-AD related cells in diseased brain so that little, if any, off-target toxicity was observed.

In parallel, our AD-correlated proteomic analysis dissected G9a-regulated pathogenesis of AD, from which proteins that showed G9a/AD-coregulated, MS1262-reversed expression or phosphorylation were mechanistically derived as new biomarkers for diagnosing AD or assessing drug effects. Particularly, in addition to multiple known AD markers that were affected by MS1262 treatment such as IL-33, complement C3, CD109, ADCYAP1, PBXIP1, CTHRC1, GFAP, OLFM3, and the protein products of 33 AD-risk genes (**Fig. 5A and Data S5C**),^106^ MS1262 treatment reversed the expression or phosphorylation of SMOC1 and other proteins in the matrisome that were recently characterized as CSF biomarkers of early stage AD ^28^ (**Figs. 6B & 7E**). Interestingly, Bellver-Sanchis et al. ^108^ showed that G9a inhibition rescued GMFB-dysregulated neuroprotection in AD. However, their transcriptomic analysis suggested that G9a is a transcriptional suppressor of *GMFB*, which failed to explain how GMFB protein was found upregulated in both AD mouse hippocampus and patient CSF (**Fig. 5B**). In agreement with our characterized function of G9a methylation of METTL3 in G9a-regulated translation ^18^, they also identified GMFB as a nonhistone substrate of G9a, which suggested similar regulation of GMFB by G9a at the translational level. Thus, our identification of AD/G9a-coupregulated, MS1262-suppressed expression of GMFB protein validated our discovery that G9a-translational mechanism outperforms G9a-suppressed transcription in defining the proteopathologic nature of AD pathogenesis.

In clinical practice, these MS1262-affected AD biomarkers can be used in combination or in a panel to 1) diagnose patients at early stage or the stages when intervention is effective, 2) evaluate evidence of drug-induced downstream disease modification, and 3) assess clinical outcomes of MS1262 treatment. Some candidate biomarkers of companion diagnosis for individualized, precision medicine were found in body fluids accessible for noninvasive assessment of drug effects. For example, although aducanumab treatment of some patients was associated with serious side effects, including brain bleeding or stroke, our mouse-to-patient conserved proteomic data revealed that MS1262 can reverse AD-characteristic expression/phosphorylation of proteins associated with blood clotting (**Fig. 6D**). These results suggested that, during therapy, MS1262 treatment may reduce or avoid the risk of blood clot burst for brain bleeding or a stroke. In addition, these Aβ abundance-correlated biomarkers represent multiple pathological brain processes that are the AD mediators of Aβ plaque, and MS1262 affected biomarkers of AD-specific Aβ pathology. Thus, these biomarkers can be used to evaluate for evidence of downstream disease modification and to assess the relationship between biomarker changes and MS1262-induced clinical outcomes.

To precisely evaluate the drug effects or individual responses to the drug, instead of conventional assays with available AD markers (if any) for one-protein-at-a-time validations, our global comparison of AD mice and AD patient proteomes^11^ determined the impact of MS1262 inhibition of G9a on AD therapy. Of clinical significance, this cross-species proteomic comparison validated the mouse-to-human conserved, G9a-translation mechanism of MS1262 reversal of AD-correlated pathways; this mouse-to-human correlation strongly suggests that MS1262 inhibition of G9a would be an effective therapy for AD patients.

In summary, for the first time, our results unambiguously indicated that defining the G9a translational mechanism is superior to characterizing the G9a epigenetic mechanism in determining the stage of AD pathogenesis and the ultimate effects of drug action. These findings also validated the mouse-to-human conserved, G9a-translational mechanism that broadly defines AD proteopathology. Accordingly, aberrantly activated G9a in diseased hippocampus regulates specific pathways at the translational or post-translational (phosphorylation) level that mechanistically contribute to the major symptoms of AD, including cognition impairment, memory loss, depression, and anxiety. Because our discovered G9a-translational mechanism directly and ultimately determined AD-correlated proteomic or phosphoproteomic changes in diseased brain, MS1262 showed high specificity and efficacy for protecting AD patients from cognitive impairment and noncognitive disorders. In addition, candidate markers of companion diagnosis can be selected from the AD-related, MS1262-reversed proteome to predict AD patient responses to MS1262 therapy and to stratify patients for enhanced therapy with high positive response rates.

## Data Availability

All data produced in the present work are contained in the manuscript

## ACKNOWLEDGMENTS

This work was supported in part by grants NIH R21AG071229, 1R41DK133051-01A1, R01 GM133107-01, and UNC University Cancer Research Fund (UCRF) (to X.C.), R01MH111773, R01MH122692, RF1AG058160, R01NS104530 (to J.S.), R01GM122749, R01HD088626 and an endowed professorship from the Icahn School of Medicine at Mount Sinai (to J.J.), NIH T32GM135095 (to R.N.S.). R01AG065611 (to Z. W.). We used Thermo-Fisher Scientific Q Exactive HF-x that was upgraded with funding from UNC Lineberger Comprehensive Cancer Center. We used the NMR Spectrometer Systems at Mount Sinai, acquired with funding from the NIH’s SIG grants 1S10OD025132 and 1S10OD028504. The indication of using MS1262 for AD therapy is protected by US provisional patent application # 63/545,673.

## AUTHOR CONTRIBUTIONS

Xian Chen conceived the project and wrote the manuscript. X. C., J.S., J.J. supervised cross-species proteomics/multiomics analysis, animal studies, MS1262 synthesis and characterization, respectively. L.X., R.N.S, and Y.X. performed and interpreted the experiments. L.X. conducted all proteomic experiments, analyzed the data, and wrote the report. A.M. analyzed proteomics and patient data and wrote the report. R.N.S and J.S. designed and performed drug treatment, analyzed animal behavior, microdissected mouse hippocampal tissues, prepared the figures, and wrote part of the manuscript. Y.X., K.S.P., J.V., J.L., and J.J. designed, synthesized, and characterized MS1262 and wrote the report. C. X. and Z. W. prepared organoids. M.D. provided APP^NLGF^ mice. Y-J.L. conducted the electrophysiology experiments and data analysis. Y-D.L. and L.Q. assisted drug treatment and mouse brain tissue dissection.

## DATA AND MATERIALS AVAILABILITY

All data needed to evaluate the conclusions in the paper are present in the paper or the Supplementary Materials. Raw files and any other data required for reanalysis are available from the corresponding author (X.C.) upon request.

## SUPPLEMENTARY MATERIALS

Figs. S1 to S6 Data File S1

## MATERIALS AND METHODS

### EXPERIMENTAL MODEL AND SUBJECT DETAILS

#### Human cerebral organoid cultures

A familial AD patient-derived iPSC line with the *APP* V717I mutation (F033K; male) and a sex- and age-matched healthy control (C-03; male) iPSC line (provided by Dr. Chadwick Hales’ laboratory at Emory University) were cultured on irradiated MEFs in human iPSC medium composed of D-MEM/F12 (Invitrogen), 20% Knockout Serum Replacement (KSR, Invitrogen), 1X Glutamax (Invitrogen), 1X MEM Non-essential Amino Acids (NEAA, Invitrogen), 100 µM β-mercaptoenthanol (Invitrogen), and 10 ng/ml human basic FGF (bFGF, PeproTech) as described ^129, 130, 131^. Forebrain-specific organoids were generated as described ^130, 131, 132^. Briefly, human iPSC colonies were detached from the feeder layer with 1 mg/ml collagenase treatment for 1 hour and suspended in embryonic body medium composed of FGF-2-free iPSC medium supplemented with 2 µM Dorsomorphin and 2 µM A-83 in nontreated polystyrene plates for 4 days with a daily medium change. On days 5-6, half of the medium was replaced with induction medium consisting of DMEM/F12, 1X N2 Supplement (Invitrogen), 10 μg/ml Heparin (Sigma), 1X Penicillin/Streptomycin, 1X Non-essential Amino Acids, 1X Glutamax, 4 ng/ml WNT-3A (R&D Systems), 1 μM CHIR99021 (Tocris), and 1 μM SB-431542 (Tocris). On day 7, organoids were embedded in Matrigel (BD Biosciences) and continued to grow in induction medium for 6 more days. On day 14, embedded organoids were mechanically dissociated from Matrigel by pipetting up and down onto the plate with a 5 ml pipette tip. Typically, 10-20 organoids were transferred to each well of a 12-well spinning bioreactor (SpinΩ) containing differentiation medium, consisting of DMEM/F12, 1X N2 and B27 Supplements (Invitrogen), 1X Penicillin/Streptomycin, 100 µM β-mercaptoenthanol (Invitrogen), 1X MEM NEAA, 2.5 μg/ml Insulin (Sigma). At day 71, differentiation medium was exchanged with maturation medium, consisting of Neurobasal (Gibco), 1X B27 Supplement, 1X Penicillin/Streptomycin, 1X β-mercaptoenthanol, 0.2 mM Ascorbic Acid, 20 ng/ml BDNF (Peprotech), 20 ng/ml GDNF (Peprotech), 1 ng/ml TFGβ (Peprotech), and 0.5 mM cAMP (Sigma). All media were changed every other day.

#### Experimental animals

All animal procedures were conducted in accordance with the NIH Guide for the Care and Use of Laboratory Animals and with the approval of the Institutional Animal Care and Use Committee at the University of North Carolina at Chapel Hill. The 5xFAD (C57BL/6J) and wild-type (C57BL/6J) littermate controls (16-24-weeks-old) were obtained from the Jackson laboratory. All 5xFAD mice were heterozygous. APP^NLGF^ (C57BL/6J-A^w-J^/J) and wild-type (C57BL/6J-A^w-J^/J) mice were obtained from Dr. Mohanish Deshmukh. All APP^NLGF^ mice were homozygous. Both male and female mice were used, and sex was matched in the various groups. No immune deficiencies or other health problems were observed in these lines, and all animals were experimentally and drug-naïve before use. Animals were group housed and bred in a dedicated husbandry facility with 12/12 hour light-dark cycles with ad libitum food and water. All mice were under veterinary supervision. Behavioral experiments were performed in the light phase. Once drug or vehicle administration began, animals were moved to a satellite housing facility with the same light-dark cycle.

### BEHAVIORAL TESTS

#### Handling

Mice were handled five days a week for 6 weeks for 5 min/day leading up to behavioral testing.

#### Open Field and Habituation

On days 1 and 2, mice were placed in an empty open field environment (45 cm square plastic chamber) for 10 minutes. After each test, the chamber was cleaned with 70% ethanol to eliminate scents from previously tested mice. The first 5 minutes of the day-1 test was analyzed by Noldus Ethovision XT to monitor animal position and locomotion. A 25 cm square was used to indicate the center of the chamber when analyzing the recordings. Both total locomotion and time spent in the center of the open field were quantified by the Noldus Ethovision XT program.

#### Novel place recognition

On day 3 of behavioral testing, also known as the encoding phase, two identical glass cylinders (height 4 cm, base diameter 1.5 cm) were fixed to the chamber floor (to prevent object movement) on the same side of the chamber 20 cm apart from each other. Animals were allowed to freely explore the chamber for 5 minutes and could interact with the objects while being recorded. Most animals showed no preference for a single object and the locations of the objects were randomized across mice. On day 4 of behavioral testing, also known as the retrieval phase, the position of one of the objects was moved to the opposite side of the chamber, and animals were recorded for 5 minutes while freely exploring the chamber and interacting with the objects. Videos were scored manually using a separately trained researcher who was blinded to the treatment group. Time spent with each object in both encoding and retrieval were scored. Any mouse that showed a preference for one object over the other (defined by interacting with one object for more than double the time of interacting with the other) during the encoding phase was not included in the analysis. Furthermore, mice that did not spend at least 2 seconds of total object interaction time (IT) were not included in the analysis. The discrimination ratio (DR) was calculated in the following way:

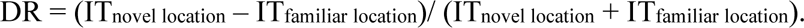

A DR of 0 indicated no preference for either object, and a DR of 0.33 indicates spending twice as much time with the object in the novel location compared with the object in the familiar location.

#### Elevated zero maze

The apparatus was an elevated white plastic ring platform (width of ring 6 cm, outer diameter 60 cm). The entire ring was elevated 60 cm off the ground. Each animal was placed in the closed arm to start the trial and was recorded for 5 minutes. Time spent in the open arm sections was scored by a trained, blinded researcher. After each trial, the apparatus was cleaned with 70% ethanol.

#### Forced Swim

The apparatus was an acrylic cylinder (diameter 20 cm, height 30 cm) filled with room temperature (23±1°C) water to a depth of 20 cm. Each mouse was recorded during a 5-minute swimming trial, and the video was later scored manually by a trained, blinded researcher for time spent immobile. Time spent immobile was defined as when all four paws of the mouse remained immobile. After each trial, the apparatus was filled with fresh water.

## METHOD DETAILS

### Drug treatment

The G9a inhibitor MS1262 was dissolved in DMSO to 10 mg/mL and aliquoted into single doses and stored at −20°C. Immediately prior to injection, these aliquots were thawed and diluted in 0.9% saline. The final solution that was injected intraperitoneally consisted of 1% DMSO and equated to a 1 mg MS1262/kg of animal weight. Control animals received 1% DMSO in 0.9% saline at weight matched volumes. Mice were randomly selected for MS1262 or vehicle treatment and received 1 injection every 3.5 days for 6 weeks.

### Microdissection of hippocampi

After 6 weeks of intermittent MS1262 or vehicle treatment, animals were anesthetized with a 5% isoflurane in oxygen mixture until the animal was no longer responsive to a toe pinch. Animals were then transcardially perfused with ice-cold PBS and the brain was isolated. The brain was sliced bilaterally across the sagittal midline. Each half was then taken, and the hippocampus was carefully dissected. Both hippocampi were placed in a cryogenic tube and flash frozen in liquid nitrogen and placed at −80°C.

### Proteomics sample preparation

The mouse hippocampus tissues were resuspended in 8 M urea, 50 mM Tris-HCl pH 8.0, reduced with dithiothreitol (5 mM final) for 30 min at room temperature, and alkylated with iodoacetamide (15 mM final) for 45 min in the dark at ambient temperature. Samples were diluted 4-fold with 25 mM Tris-HCl pH 8.0, 1 mM CaCl_2_ and digested with trypsin at a ratio of 1:100 (w/w, trypsin: protein) overnight at ambient temperature. There were three wild-type (WT) controls, 4 AD, and 4 AD-treated with MS1262. Peptides were cleaned by homemade C18 stage tips and the concentration was determined (Peptide assay, Thermo 23275). One hundred microgram each was used for labeling with isobaric stable tandem mass tags (TMT^11^, Thermo Fisher Scientific, San Jose, CA) following supplier instruction. The mixture of labeled peptides was desalted on Cep-Pak light C18 cartridge (Waters). Phosphopeptides were enriched with High-Select Fe-NTA Phosphopeptide Enrichment Kit (Thermo Scientific). One hundred microgram of peptides was fractionated into 20 fractions on C18 stage tip with 10 mM trimethylammonium bicarbonate (TMAB), pH 8.5 containing 5 to 50% acetonitrile.

### Mass spectrometry

Dried peptides were dissolved in 0.1% formic acid, 2% acetonitrile. One microgram phosphopeptides or 0.5 μg of peptides from each fraction was analyzed on a Q-Exactive HF-X coupled with an Easy nanoLC 1200 (Thermo Fisher Scientific, San Jose, CA). Peptides were loaded on to a nanoEase MZ HSS T3 Column (100Å, 1.8 µm, 75 µm x 250 mm, Waters). The phosphopeptides were separated with 240-min gradient: a linear gradient of 5 to 20% buffer B over 140 min, 20 to 31% over 50 min, 31 to 75% over 30 min, followed by a ramp to 100% B in 1 min and 19-min wash with 100% B. Analytical separation of unphosphorylated peptides was achieved with 110-min gradient, a linear gradient of 5 to 10% buffer B over 5 min, 10% to 31% buffer B over 70 min, 31% to 75% buffer B over 15 followed a ramp to 100%B in 1 min and 19-min wash with 100%B, where buffer A was aqueous 0.1% formic acid, and buffer B was 80% acetonitrile and 0.1% formic acid. The flow rate was kept at a 250 nl/min. Mass spectrometry experiments were also conducted in a data-dependent mode with full MS (externally calibrated to a resolution of 60,000 at *m*/*z* 200) followed by high energy collision-activated dissociation-MS/MS of the top 10 most intense ions with a resolution of 45,000 at *m/z* 200. High energy collision-activated dissociation-MS/MS was used to dissociate peptides at a normalized collision energy of 32 eV in the presence of nitrogen bath gas atoms. Dynamic exclusion was 45 seconds.

### Raw proteomics data processing and analysis

Peptide identification and quantification with TMT reporter ions were performed using the MaxQuant software version 2.1.0.0 (Max Planck Institute, Germany). Protein database searches were performed against the UniProt human protein sequence database (UP000005640). A false discovery rate for both peptide-spectrum match and protein assignment was set at 1%. Search parameters included up to two missed cleavages at Lys/Arg on the sequence, oxidation of methionine, protein N-terminal acetylation, and phosphorylation of serine, threonine, and tyrosine as dynamic modifications. Carbamidomethylation of cysteine residues was considered as a static modification. Peptide identifications are reported by filtering of reverse and contaminant entries and assigning to their leading razor protein. Data processing and statistical analysis were performed on Perseus (Version 1.6.10.50). Protein quantitation was performed on biological replicates and a two-sample t-test statistics was used with a p-value of 5% to report statistically significant protein or phosphopeptide abundance fold-changes.

### Slice preparation

Acute slices were prepared from 5.25 month-5.5 month male 5xFAD mice treated with MS1262 (1 mg/kg) or vehicle as described previously. Animals were anesthetized with isoflurane (5% in O_2_) and transcardially perfused with ice-cold oxygenated artificial cerebrospinal fluid (ACSF) containing the following items (in mM): 92 NMDG, 30 NaHCO_3_, 25 glucose, 20 HEPES, 10 MgSO_4_, 5 sodium ascorbate, 3 sodium pyruvate, 2.5 KCl, 2 thiourea, 1.25 NaH_2_PO_4_, and 0.5 CaCl_2_ (pH 7.3, 310 mOsm). Brains were rapidly removed, and acute transverse hippocampal slices (280 μm) were cut using a Leica vibratome (VT1200, Germany). Next, slices were warmed to 34.5°C for 8 minutes. Then, slices were maintained in the holding chamber containing HEPES ACSF (in mM): 92 NaCl, 30 NaHCO_3_, 25 glucose, 20 HEPES, 5 sodium ascorbate, 3 sodium pyruvate, 2.5 KCl, 2 thiourea, 2 MgSO_4_, 2 CaCl_2_, 1.25 NaH_2_PO_4_ (pH 7.3, 10 mOsm) at ambient temperature for at least 1 hour before recording. Electrophysiological recordings were made at 32°C using a heater controller (TC-324C, Warner Instruments) in ACSF containing (in mM): 125 NaCl, 26 NaHCO_3_, 20 glucose, 2.5 KCl, 2 CaCl_2_, 1.3 MgSO_4_, 1.25 NaH_2_PO_4_, (pH 7.3, 310 mOsm). The flow rate was 2 ml/min.

### Electrophysiology

Slices were visualized on a fixed-stage upright microscope (Olympus BX51WI) equipped with ×4 and ×40 objectives and differential interference contrast optics, infrared illumination, and an infrared-sensitive camera. Whole-cell patch-clamp recordings were performed with glass pipettes with resistance of 4.0–6.0 MΩ when filled with internal solution containing (in mM): 130 K-gluconate, 20 HEPES, 4 MgCl_2_, 4 Na-ATP, 2 NaCl, 0.5 EGTA, 0.4 Na-GTP (pH 7.2, 290 mOsm). Series resistance (Rs) was monitored throughout all experiments, and cells with Rs changes over 20% were discarded. All recordings were made from granule cells located in the middle or outer layer of the dentate gyrus. For spontaneous excitatory postsynaptic currents (sEPSCs) recordings, dentate granule cells were held at −70 mV in voltage-clamp mode and bicuculine (20 μM; GABA_A_ receptor antagonist; Tocris Bioscience) was added to the ACSF. Intrinsic cellular properties were recorded in current-clamp mode. To test neuronal input resistance, hyperpolarizing current pulses (20 pA, 200 ms) were applied to the neurons. Resting membrane potential was measured as the membrane potential baseline value obtained in current-clamp mode in the absence of current injection. The current-voltage relationship experiments (to evaluate action potential firing rate) consisted of a series of current injections (500-ms duration) between 0 pA and +240 pA in 20 pA delivered in step-wise increments. Data were acquired with a Multiclamp 700B amplifier and digitized with a Digidata 1440A using pCLAMP 10.7 acquisition software (Molecular Devices).

## Supplementary Materials

**Other Supplementary Materials for this manuscript include the following:**

Data File S1

**Data File S1. (Table 1)** List of ChaC-MS identifications of UNC0965 pulled down proteins from human organoids.

**Data File S1. (Table 2)** List of ChaC-MS identifications of UNC0965 pulled down proteins from the hippocampus of wild type or 5xFAD mice with or without MS1262 treatment, which are labeled as ‘WT’, ‘AD’, ‘AD-tr’ in the Table.

**Data File S1. (Table 3)** List of protein and phosphoprotein identifications in the hippocampus of age matched wildtype controls (WT) or 5xFAD/APP-NLFG mice (AD) or AD mice with MS1262-treatment (AD-tr).

**Data File S1. (Table 4)** Tables summarizing activity scores for overrepresented pathways and diseases & biological functions calculated using differentially expressed proteins/phosphoproteins listed in Table 3

**Data File S1. (Table 5)** AD/G9a-corregulated proteins/phosphor-proteins whose expression shows significant correlation with Abeta peptide (6-28) in the hippocampus of AD patients.

---

**Figure S1.**
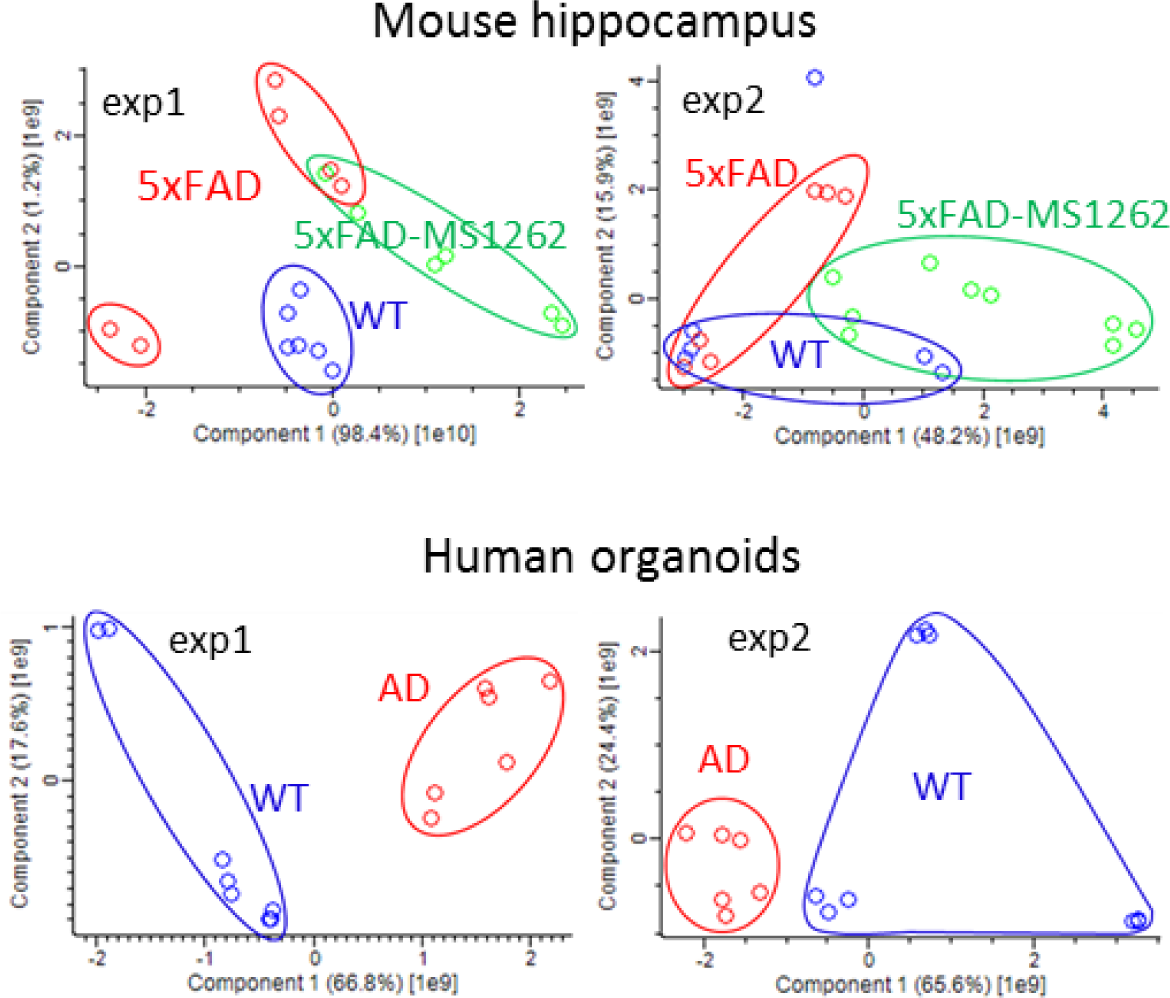
PCA plots of ChaC pull-downs by UNC0965 from mouse hippocampus and human organoids.

**Figure S2.**
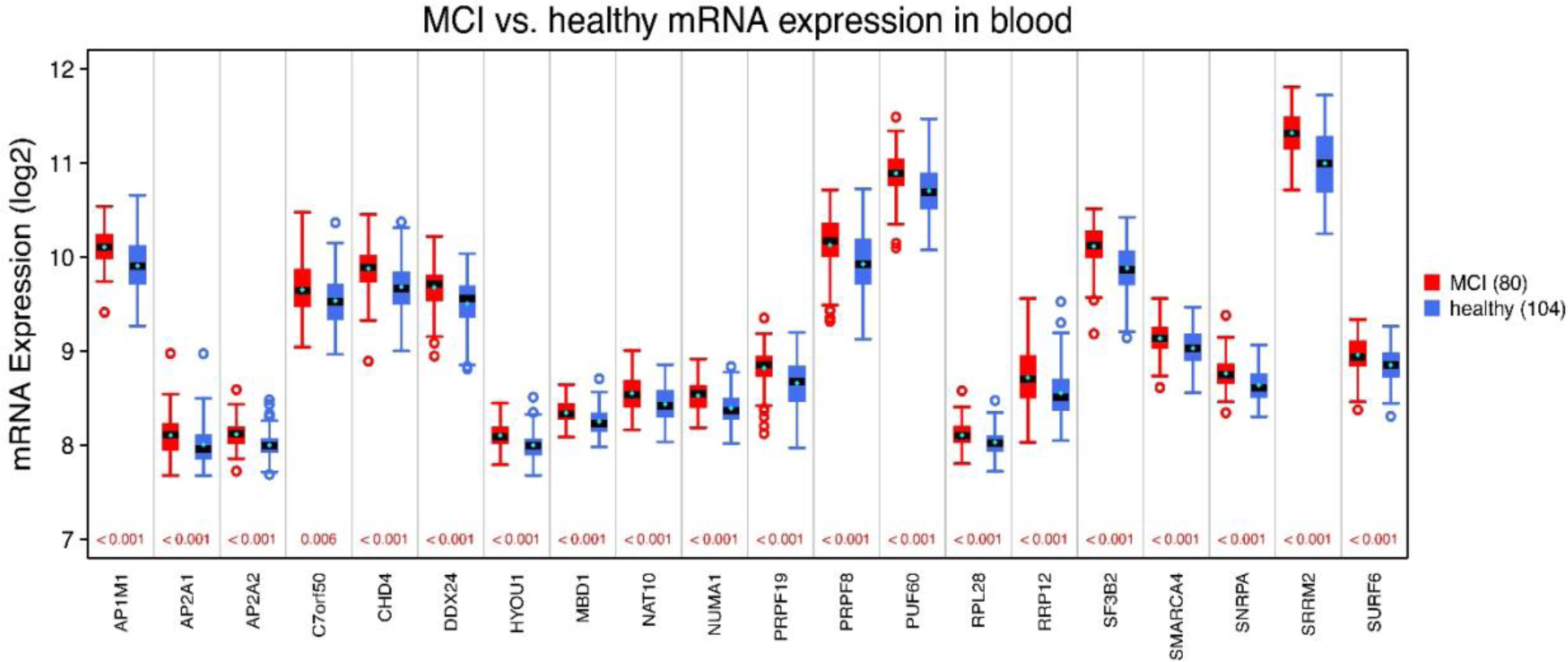
ChaC-identified G9a interactors show the interaction-correlated overexpression patterns in MCI or AD patients. Box plots showing the statistically significant (p values < 0.05), patient-specifically altered mRNA expression for indicated interactor genes. The distribution of mRNA expression for MCI patients (n=80) is in red, and for healthy individuals (n=104) is in blue. Standard error of the mean (SEM) in cyan is in ± 0.01-0.04.

**Figure S3.**
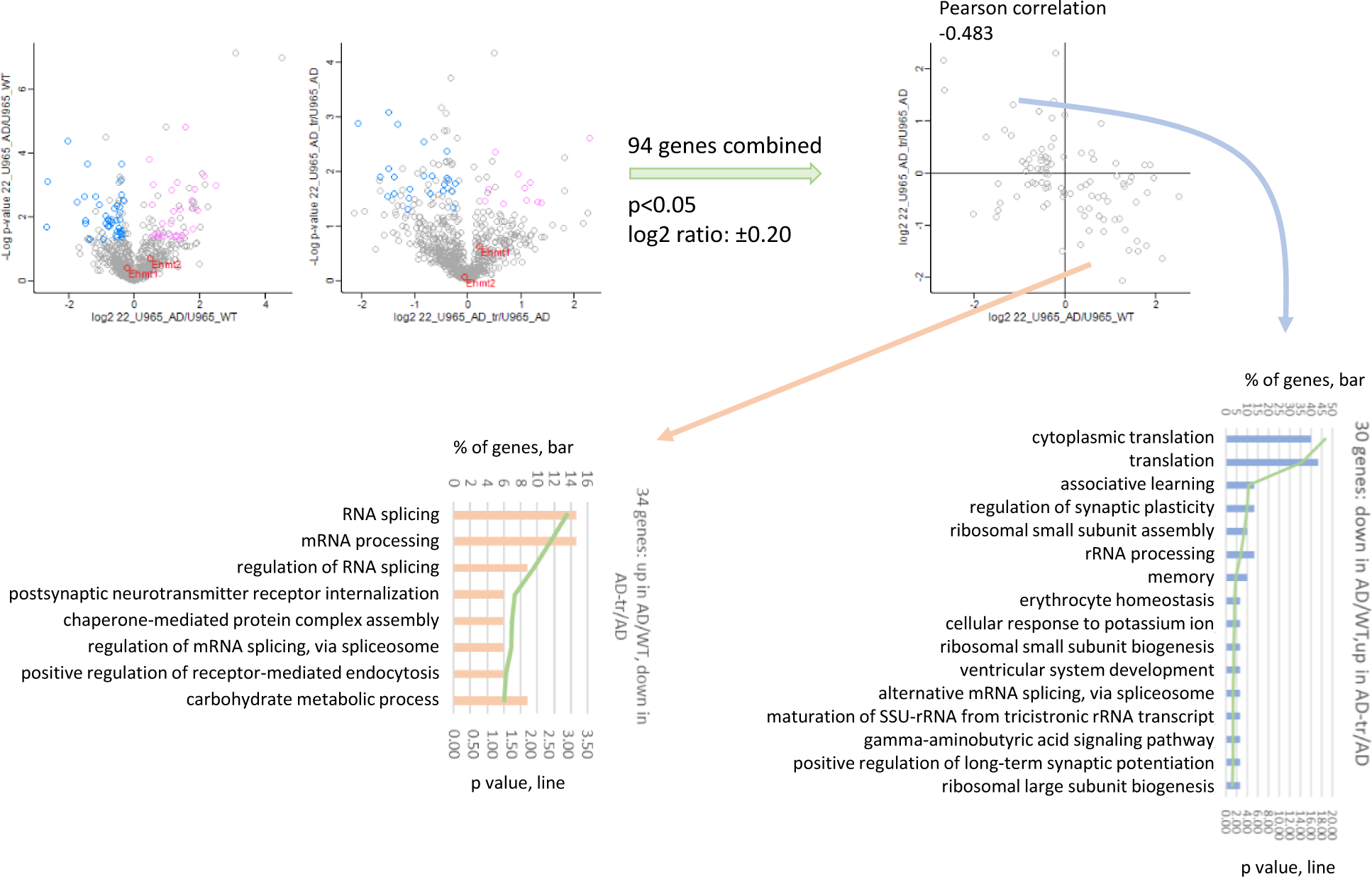
Function categorization of ChaC-MS identifications of G9a interactors from the hippocampus of 5XFAD mice with versus without MS1262 treatment.

**Figure S4.**
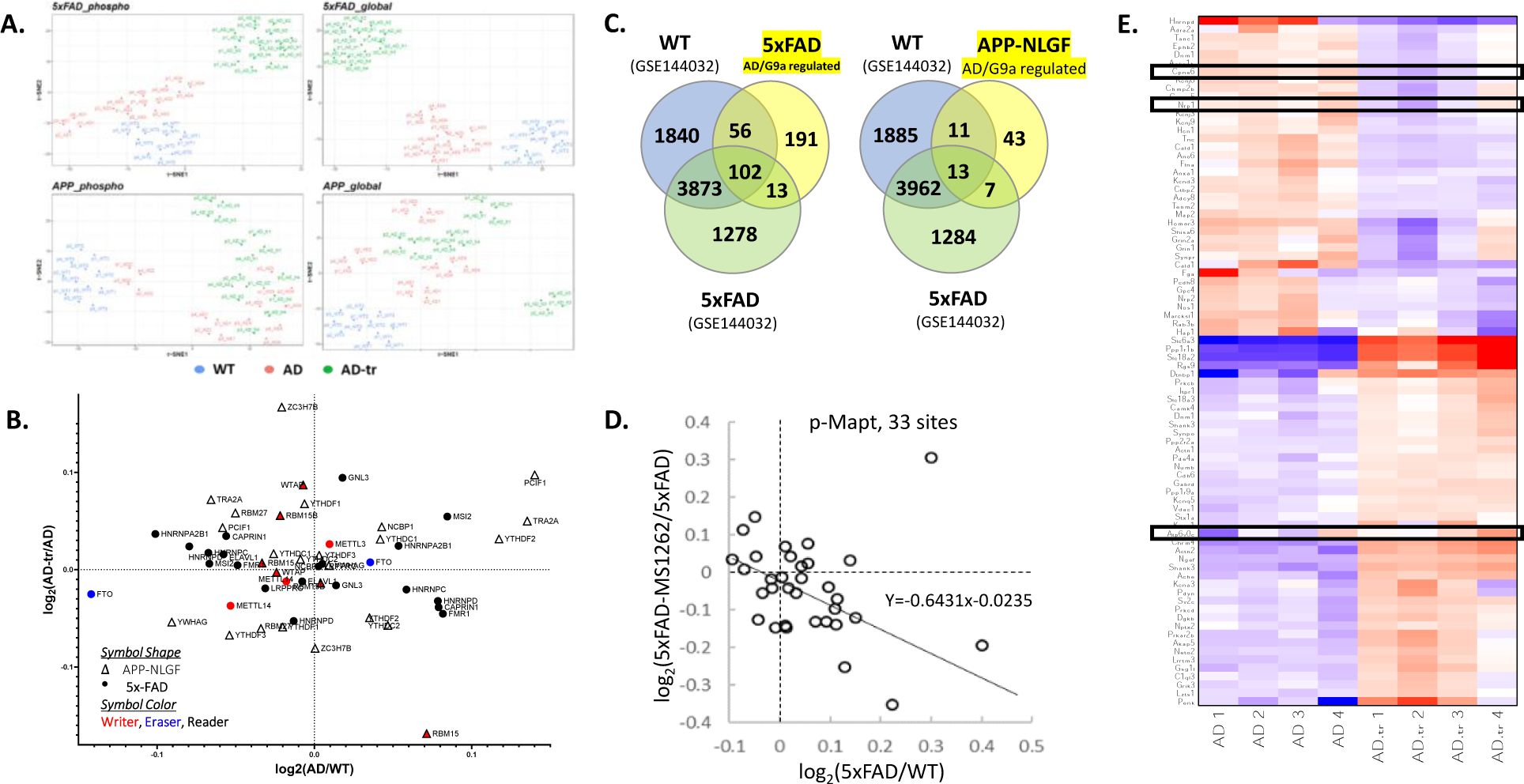
**(A)** t-SNE visualization showing clear separation between and reproducibility among replicates in the proteomic and phospho-proteomic datasets. ‘WT’– age-matched wild type mice; ‘AD’– 5x-FAD/APP-NLGF mice at mid/late stages of AD; ‘AD-tr’ – the AD mice treated by MS1262. **(B)** Scatter plot showing difference in expression for various m6A readers (black), writers (red) and erasers (blue) for indicated comparisons: AD vs WT (x-axis) and AD-tr vs AD (y-axis). Global proteomics data from 5x-FAD (circles) and APP-NLGF (triangles) mice. **(C)** Venn diagram showing overlap among AD-dysregulated/MS1262-reversed (‘AD/G9a coregulated’) proteins from the two mouse models (this study) and m6A-modified transcripts in WT and 5x-FAD mice identified using m6A-Seq (GSE144032). **(D)** Scatter plot showing effect of MS1262 treatment on phosphorylation of 33 sites on MAPT protein in 5x-FAD mice. **(E)** Heatmap of select AD/G9a coregulated proteins of interest. Median of 4 replicates is plotted.

**Figure S5.**
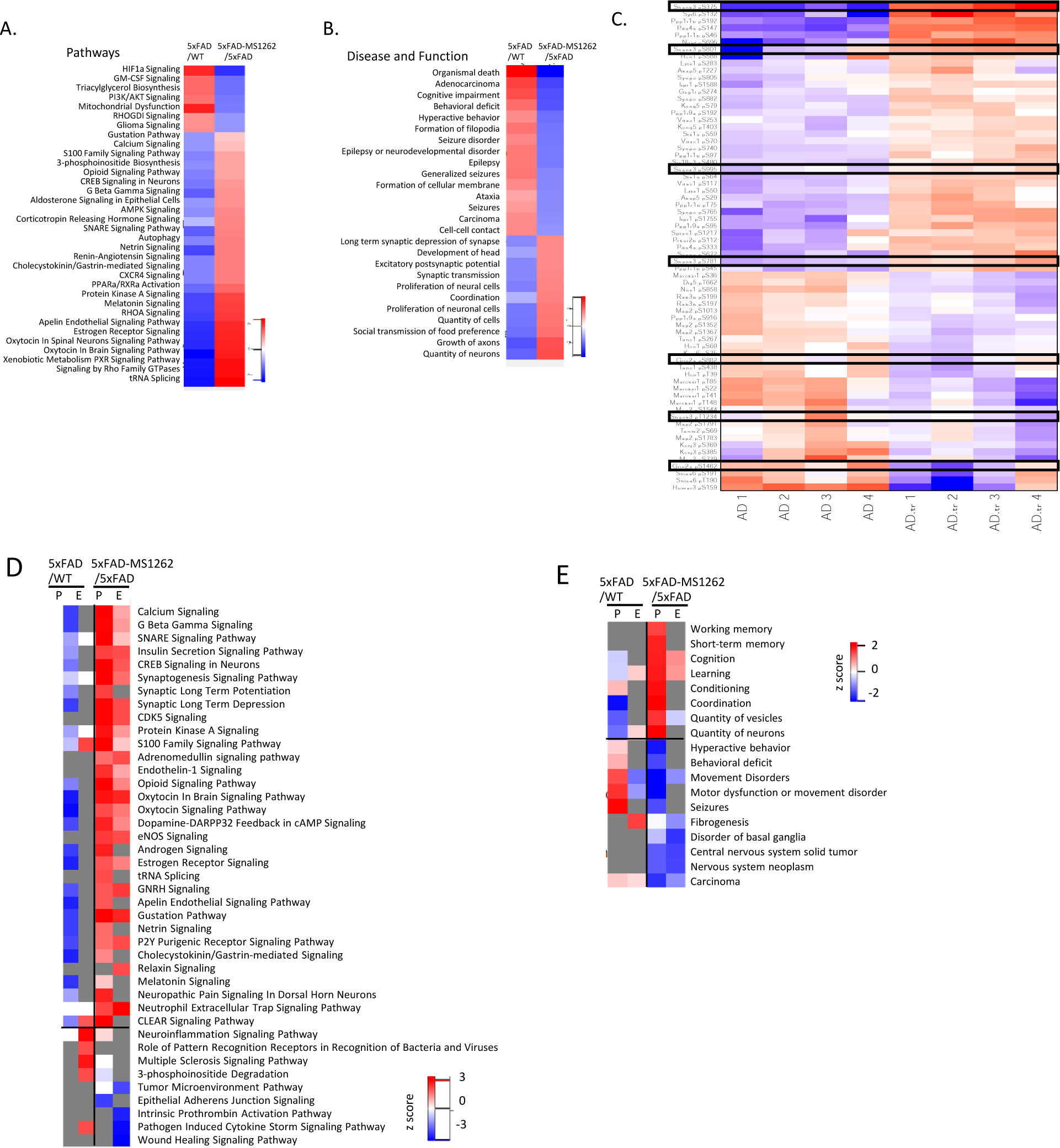
Effect of MS1262 treatment in 5xFAD mice. (**A-B**) Heatmaps summarizing pathway enrichment (A) & disease and function (B) activity analysis results calculated using differentially regulated phosphoproteins for the indicated comparisons (i.e., 5xFAD-vs-WT and 5xFAD-MS1262 vs 5xFAD). **(C)** Heatmap showing MS1262-regulated phosphoprotein products for AD-risk genes (p value < 0.05) (Nature genetics 51.3 (2019): 414-430). Midian of 4 technical replicates is plotted for each biological replicate. (**D-E**) Heatmaps summarizing pathway enrichment (A) & disease and function (B) activity analysis results calculated using differentially regulated proteins (Expression, ‘E’) & phosphoproteins (‘p’) for the indicated comparisons (i.e., 5xFAD-vs-WT and 5xFAD-MS1262 vs 5xFAD).

**Figure S6.**
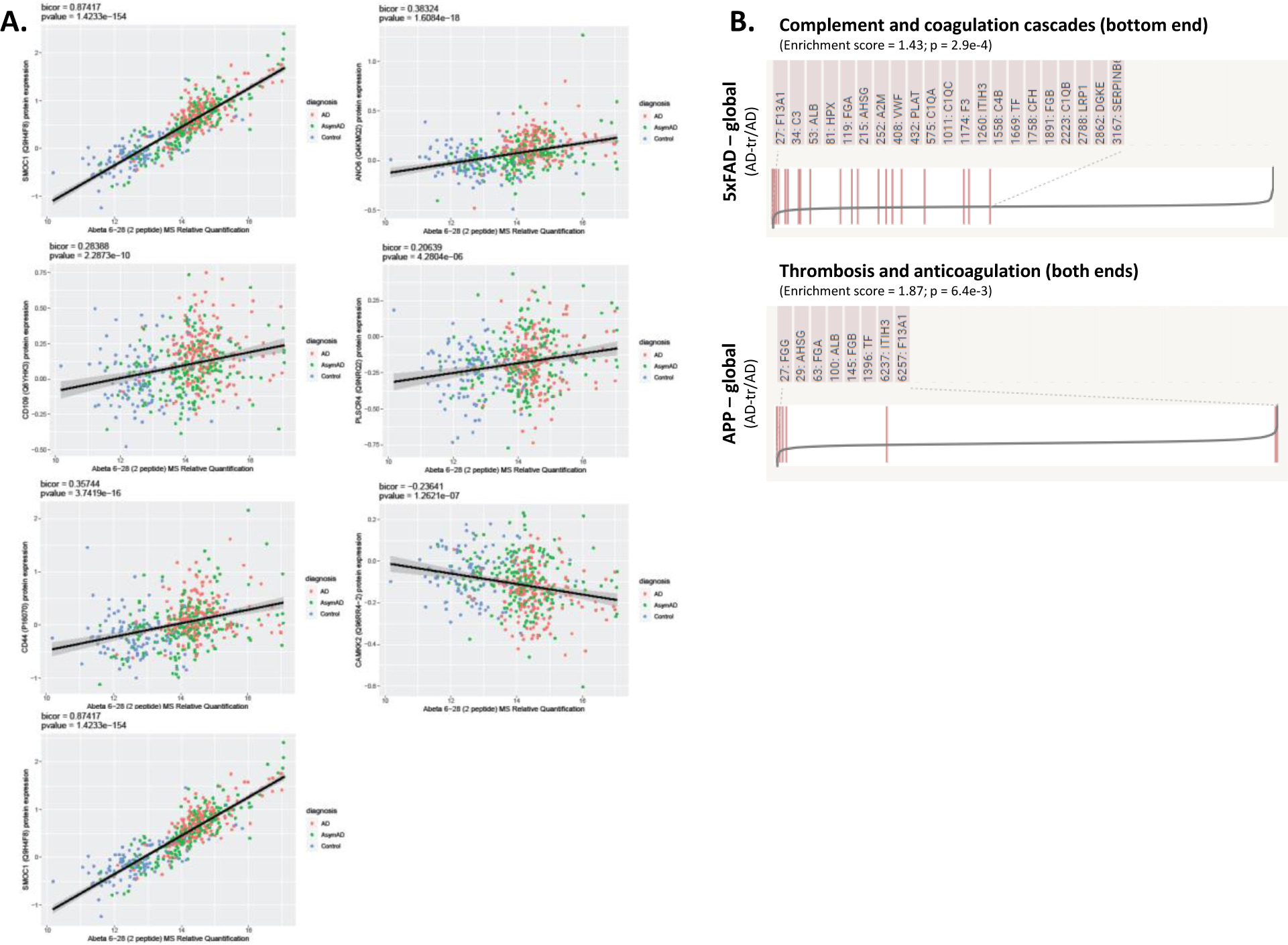
(A) Correlation plots between expression of indicated proteins and Aβ peptides (residues 6-28) in the hippocampus of AD patients (n = 488; Nature Neuroscience 25, 213-225 (2022)). Linear regression line with 95% CI is shown. Points are colored based on diagnosis of individual patients: ‘AD’ - Symptomatic AD patient; ‘AsymAD’ - asymptomatic AD patient; ‘Control’ control patients. The biweight midcorrelation (bicor) and corresponding p-value is shown on top and was calculated using the bicorAndPvalue function from the WGCNA R package. **(B)** MS1262 treatment leads to downregulation of coagulation related factors in both 5x-FAD and APP-NLGF mice.

